# GestaltMatcher Database - A global reference for facial phenotypic variability in rare human diseases

**DOI:** 10.1101/2023.06.06.23290887

**Authors:** Hellen Lesmann, Alexander Hustinx, Shahida Moosa, Hannah Klinkhammer, Elaine Marchi, Pilar Caro, Ibrahim M. Abdelrazek, Jean Tori Pantel, Merle ten Hagen, Meow-Keong Thong, Rifhan Azwani Binti Mazlan, Sok Kun Tae, Tom Kamphans, Wolfgang Meiswinkel, Jing-Mei Li, Behnam Javanmardi, Alexej Knaus, Annette Uwineza, Cordula Knopp, Tinatin Tkemaladze, Miriam Elbracht, Larissa Mattern, Rami Abou Jamra, Clara Velmans, Vincent Strehlow, Maureen Jacob, Angela Peron, Cristina Dias, Beatriz Carvalho Nunes, Thainá Vilella, Isabel Furquim Pinheiro, Chong Ae Kim, Maria Isabel Melaragno, Hannah Weiland, Sophia Kaptain, Karolina Chwiałkowska, Miroslaw Kwasniewski, Ramy Saad, Sarah Wiethoff, Himanshu Goel, Clara Tang, Anna Hau, Tahsin Stefan Barakat, Przemysław Panek, Amira Nabil, Julia Suh, Frederik Braun, Israel Gomy, Luisa Averdunk, Ekanem Ekure, Gaber Bergant, Borut Peterlin, Claudio Graziano, Nagwa Gaboon, Moisés Fiesco-Roa, Alessandro Mauro Spinelli, Nina-Maria Wilpert, Prasit Phowthongkum, Nergis Güzel, Tobias B. Haack, Rana Bitar, Andreas Tzschach, Agusti Rodriguez-Palmero, Theresa Brunet, Sabine Rudnik-Schöneborn, Silvina Noemi Contreras-Capetillo, Ava Oberlack, Carole Samango-Sprouse, Teresa Sadeghin, Margaret Olaya, Konrad Platzer, Artem Borovikov, Franziska Schnabel, Lara Heuft, Vera Herrmann, Renske Oegema, Nour Elkhateeb, Sheetal Kumar, Katalin Komlosi, Khoushoua Mohamed, Silvia Kalantari, Fabio Sirchia, Antonio F. Martinez-Monseny, Matthias Höller, Louiza Toutouna, Amal Mohamed, Amaia Lasa-Aranzasti, John A. Sayer, Nadja Ehmke, Magdalena Danyel, Henrike Sczakiel, Sarina Schwartzmann, Felix Boschann, Max Zhao, Ronja Adam, Lara Einicke, Denise Horn, Kee Seang Chew, KAM Choy Chen, Miray Karakoyun, Ben Pode-Shakked, Aviva Eliyahu, Rachel Rock, Teresa Carrion, Odelia Chorin, Yuri A. Zarate, Marcelo Martinez Conti, Mert Karakaya, Moon Ley Tung, Bharatendu Chandra, Arjan Bouman, Aime Lumaka, Naveed Wasif, Marwan Shinawi, Patrick R. Blackburn, Tianyun Wang, Tim Niehues, Axel Schmidt, Regina Rita Roth, Dagmar Wieczorek, Ping Hu, Rebekah L. Waikel, Suzanna E. Ledgister Hanchard, Gehad Elmakkawy, Sylvia Safwat, Frédéric Ebstein, Elke Krüger, Sébastien Küry, Stéphane Bézieau, Annabelle Arlt, Eric Olinger, Felix Marbach, Dong Li, Lucie Dupuis, Roberto Mendoza-Londono, Sofia Douzgou Houge, Denisa Weis, Brian Hon-Yin Chung, Christopher C.Y. Mak, Hülya Kayserili, Nursel Elcioglu, Ayca Aykut, Peli Özlem Şimşek-Kiper, Nina Bögershausen, Bernd Wollnik, Heidi Beate Bentzen, Ingo Kurth, Christian Netzer, Aleksandra Jezela-Stanek, Koen Devriendt, Karen W. Gripp, Martin Mücke, Alain Verloes, Christian P. Schaaf, Christoffer Nellåker, Benjamin D. Solomon, Markus M. Nöthen, Ebtesam Abdalla, Gholson J. Lyon, Peter M. Krawitz, Tzung-Chien Hsieh

## Abstract

The most important factor that complicates the work of dysmorphologists is the significant phenotypic variability of the human face. Next-Generation Phenotyping (NGP) tools that assist clinicians with recognizing characteristic syndromic patterns are particularly challenged when confronted with patients from populations different from their training data. To that end, we systematically analyzed the impact of genetic ancestry on facial dysmorphism. For that purpose, we established the GestaltMatcher Database (GMDB) as a reference dataset for medical images of patients with rare genetic disorders from around the world. We collected 10,980 frontal facial images – more than a quarter previously unpublished - from 8,346 patients, representing 581 rare disorders. Although the predominant ancestry is still European (67%), data from underrepresented populations have been increased considerably via global collaborations (19% Asian and 7% African). This includes previously unpublished reports for more than 40% of the African patients. The NGP analysis on this diverse dataset revealed characteristic performance differences depending on the composition of training and test sets corresponding to genetic relatedness. For clinical use of NGP, incorporating non-European patients resulted in a profound enhancement of GestaltMatcher performance. The top-5 accuracy rate increased by +11.29%. Importantly, this improvement in delineating the correct disorder from a facial portrait was achieved without decreasing the performance on European patients. By design, GMDB complies with the FAIR principles by rendering the curated medical data findable, accessible, interoperable, and reusable. This means GMDB can also serve as data for training and benchmarking. In summary, our study on facial dysmorphism on a global sample revealed a considerable cross ancestral phenotypic variability confounding NGP that should be counteracted by international efforts for increasing data diversity. GMDB will serve as a vital reference database for clinicians and a transparent training set for advancing NGP technology.

## Introduction

Facial dysmorphism is one of the most complex and informative clinical features in syndromic disorders, and is therefore often crucial in terms of establishing a diagnosis in rare genetic diseases^1,2^. However, the recognition of dysmorphic patterns, is a challenging endeavour, and relies on the skills, knowledge, and experience of the examiner. In certain syndromes, in particular those that are ultra-rare, variability in facial features can pose challenges even for highly experienced clinicians^3^. Facial features can also vary according to sex, age, and ancestry, which further complicates the recognition of a specific dysmorphic pattern^4–6^.

Ancestry plays a particularly significant role since considerable inter-ancestral variability exists in facial gestalt^7^. Thus, facial features that are common in certain ancestral groups may be considered dysmorphic in others. For example, while upslanting palpebral fissures are common in healthy Asians, they may be perceived as dysmorphic in other populations^8^. Previous studies have also highlighted differences in facial gestalt between different ancestries in common dysmorphic genetic syndromes such as Down Syndrome, 22q11.2 deletion syndrome, Noonan syndrome, and Williams–Beuren syndrome^4,9,10^. Furthermore, Lumaka et al. have demonstrated that this variability can influence the assessor, with European clinicians failing to recognize dysmorphic features in individuals of African ancestry^11^. This is a growing problem as globalization and migration increasingly blur ancestral and cultural boundaries, and geography is no longer a key determining factor in mating patterns^12^. Hence, in diverse populations, such as those with admixed ancestries, the challenge of accurately diagnosing rare diseases becomes even more pronounced since new phenotypes can evolve via admixture^13^.

Ancestry also has a significant impact on the detection of rare dysmorphic disorders via artificial intelligence (AI)^11^ because in most healthcare datasets, non-European ancestries are underrepresented^14^. Many next-generation phenotyping (NGP) approaches that predict disorders on the basis of facial image analysis, such as GestaltMatcher^15^, have demonstrated high accuracy in patients from the ancestries in which they were predominantly trained and validated, i.e., European and North American^15–19^.

Since the significantly higher birth rates in non-European regions account for 80% of the global population and 90% of all annual births (Figure 1a)^20^, action is required to include non-European patients currently considered to be underrepresented. So far, few studies exist about the performance of NGP tools where the ancestry composition of individuals in the training and test set differs. Literature suggests that AIs trained on individuals of European ancestry perform better on a test set of Asian rather than African ancestry^21–24^ that may be explained by their closer genetic relatedness^25^. This raises the question of whether AIs need to be trained for different ancestries or whether a similar performance can be achieved by sufficiently increasing the ancestral diversity in the joint training set. The latter is indicated by a study conducted on Down syndrome patients of African ancestry^11^. However, comparing these studies is difficult since they were not performed on data compliant with FAIR principles that are findable, accessible, interoperable, and reusable, meaning the results cannot be reproduced.

**Figure 1.**
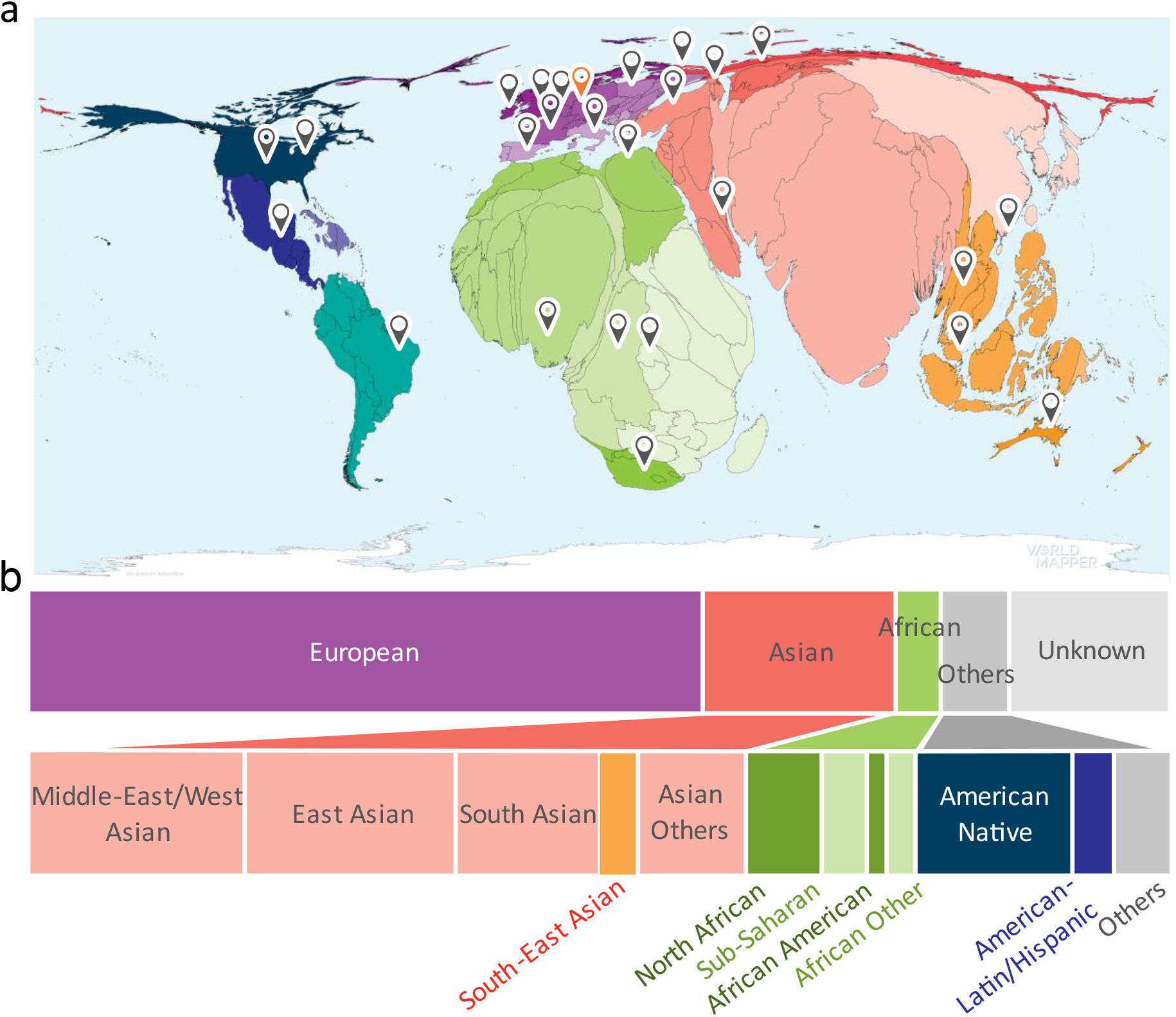
**a)** Birth rate distribution worldwide. The size of country is scaled in accordance with the respective birth rate. The map indicates countries from which unpublished images were obtained (source: https://worldmapper.org/faq/, modified). **b)** Distribution of ancestry groups in GestaltMatcher Database. 16% of the patients without ancestral information were categorized as Unknown. The breakdown of ancestries in the dataset with known ancestry is as follows: European 67%, Asian 19%, African 7%, and Others 7%.

The motivation of our work is therefore threefold: 1) scientific, because we wanted to study the effect of inter- and intra-ancestral phenotypic variability on NGP, such as GestaltMatcher, in a systematic manner; 2) clinical, because more diverse training data can presumably increase the performance of NGP on non-European ancestries; and 3) societal, because so far underrepresented populations would benefit from potential performance improvements.

To achieve these goals, we aimed for a FAIR database with an increased number of patients of non-European ancestry with respect to comparable databases^20,26,27^. Therefore, we established the GestaltMatcher Database (GMDB) as a community-driven online framework that facilitates acquiring patient consent and incentivizes data sharing, acknowledging contributions from clinician-scientists as citeable micro-publications (Figure 2)^28–31^. Through this framework, we established global collaborations, enabling the collection of a wide range of data from various ancestries.

**Figure 2.**
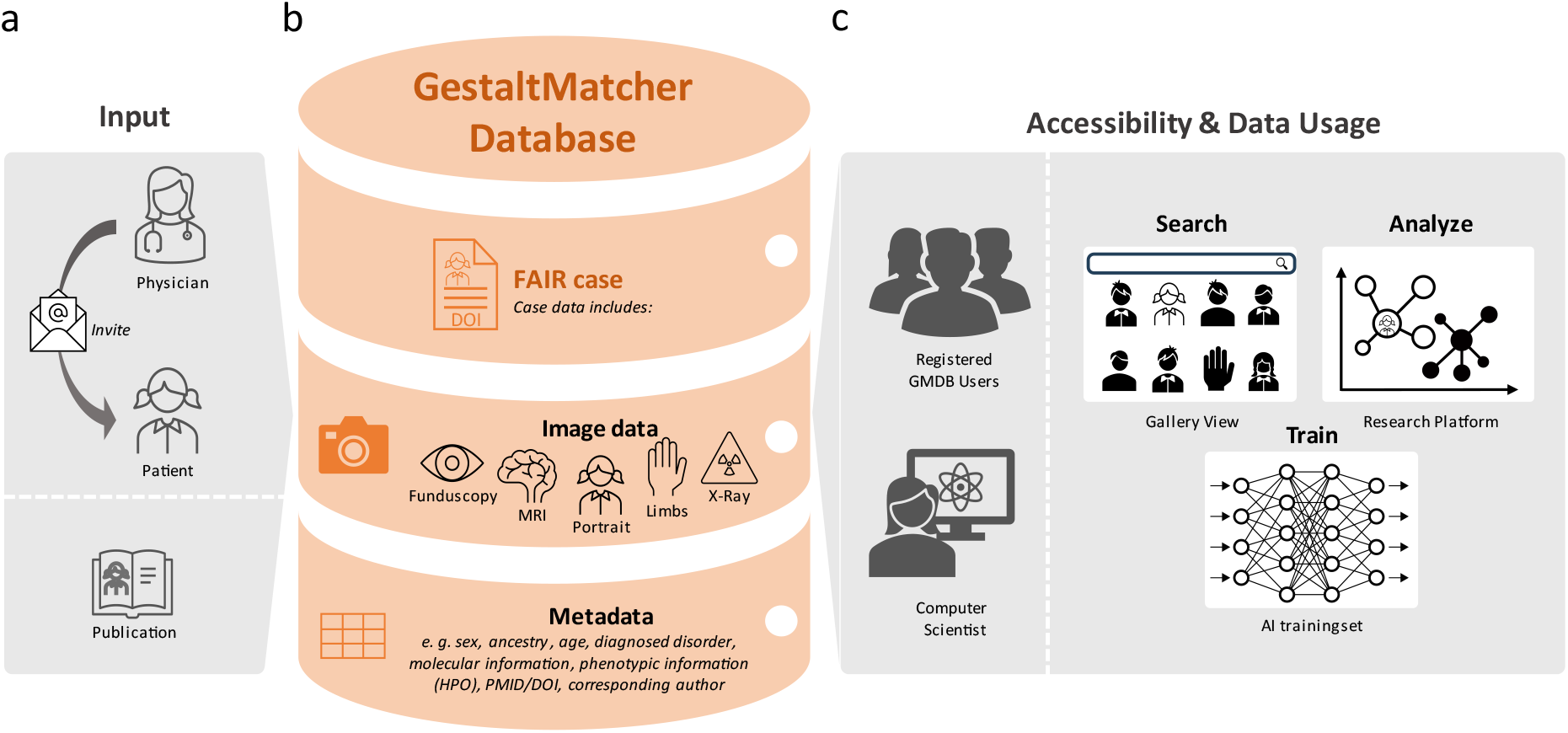
GestaltMatcher Database (GMDB) Architecture and Dataflow. **a)** Retrospective data are collected from the literature and annotated by data curators or are uploaded by collaborating attending clinician. Patients can also upload images of their own cases, incorporate prospective data, and view their own data at any time. **b)** The data (multimodal image data, including portrait images as well as magnetic resonance imaging, X-ray, fundscopy and extremity images) are stored in the GMDB (MySQL database) together with the relevant meta information (such as sex, age, ancestry, molecular, and phenotypic information). **c)** Registered users can view and search the FAIR data in the GMDB Gallery. The patient image can also be analyzed using the Next-Generation Phenotyping tool GestaltMatcher within the Research Platform. In addition, once their application has been approved by the Advisory Board, external computer scientists can use the GMDB-FAIR data set for training purposes for their projects.

GMDB is the first database for medical imaging data of patients with rare genetic disorders from diverse ancestries that is compliant with the FAIR principles^32^. By its machine-readable design, GMDB also enables systematic analyses of the influence of genetic background on NGP performance, which we will report in this study.

## Results

### Overview of FAIR data in GMDB

Retrospective data from curated publications, along with data provided by clinicians or patients, were made available as FAIR cases in the GMDB (Figure 3, Supplementary Figures 1 and 2)^33^. At the time of the data freeze for this paper on April 6^th^ 2024, we curated the GMDB-FAIR dataset consisting of 10,980 portrait images (Supplementary Figure 3) of 8,346 patients with 581 genetic disorders, including patients curated from 2,224 scientific publications. 2,312 unpublished images were contributed by 138 clinicians from 106 institutions (indicated by location markers in Figure 1a), including novel cases from GMDB micro-publications (micro-publication section in Supplementary Note). For the portrait data, which is the scope of this study, in terms of sex, the data distribution is relatively balanced (Figure 4a). However, age is biased toward patients aged below 10 years (Figure 4b). Figure 4c shows a two-dimensional representation of Human Phenotype Ontology^34^ (HPO)-defined symptom groups in GMDB via Uniform Manifold Approximation and Projection (UMAP). While GMDB incorporates cases from all HPO-defined symptom groups across the disease landscape, the HPO-defined symptom group ‘facial dysmorphism’ is enriched in GMDB. Since each individual can be attributed to several HPO-defined symptom groups according to their features, facial dysmorphism was also present in the other HPO-defined symptom groups, as shown in the heatmap.

**Figure 3.** An example case presentation of a FAIR case with a Digital Object Identifier (DOI). **a)** FAIR cases in the GestaltMatcher Database (GMDB) are displayed to GMDB users via the data sheet. Each FAIR case can also be assigned a DOI in order to render it a citable micro-publication. This micro-publication contains the image data and metadata, including demographic, molecular, and phenotype information. The dynamic nature of the GMDB case report enables longitudinal image data storage even after initial publication, which is not possible in conventional journals. **b)** After uploading, case reports can be viewed and searched by other users in the Gallery view. **c)** The image data can also be used for inter-cohort comparisons of the gestalt scores within the research platform.

**Figure 4.**
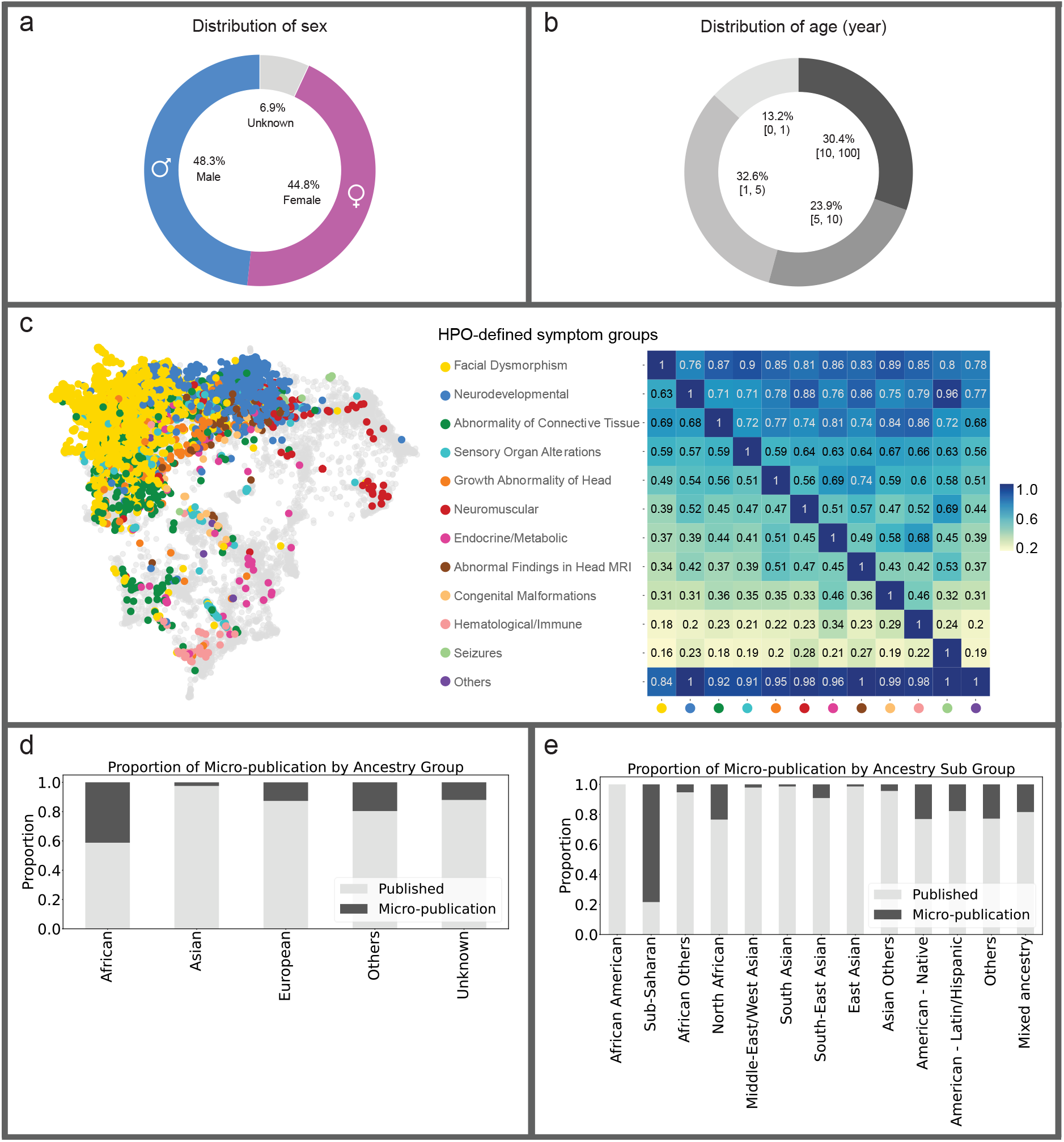
Overview of the GestaltMatcher Database (GMDB)-FAIR dataset. **a)** Sex distribution. Number of images shown in brackets. **b)** Distribution of patient age in years. **c)** Left: Two-dimensional representation of phenotypic similarities between patients, as calculated on the basis of Human Phenotype Ontology (HPO) terms via Uniform Manifold Approximation and Projection (UMAP). HPO terms were annotated for 4,474 individuals in the GMDB, and expert clinicians defined twelve distinct HPO-defined symptom groups. Based on the annotated HPO terms, each case was assigned to one or more HPO-defined symptom groups. All OMIM diseases were included, using their HPO annotations (gray background dots) as a reference. GMDB cases are color-coded according to their most pronounced HPO-defined symptom group, i.e., the group that includes the majority of their HPO terms. The dataset is dominated by two major clusters (facial dysmorphism in yellow and neurodevelopmental in blue) but shows cases from across the complete disease landscape. Right: Heatmap of the proportion of GMDB individuals within the HPO-defined symptom group on the X-axis who are also assigned to the HPO-defined symptom group on the Y-axis. Notably, facial dysmorphism is present in at least 70% of the cases of each HPO-defined symptom group. **d)** Proportion of the unpublished and published images in each ancestry group. **e)** Proportion of the unpublished and published images in each sub-ancestry group.

### Underrepresented populations benefited from micro-publication case reports in GMDB

Through our international collaborations (Figure 1a), the representation of non-European ancestral groups is 19% for Asian, 7% for African, and 7% for Others. 67% comprises individuals of European descent (Figure 1b). Moreover, the ancestry distribution varies among different disorders. Some disorders, such as Williams-Beuren syndrome, Hyperphosphatasia with impaired intellectual development syndrome, and Cohen syndrome, have relatively diverse and balanced ancestral distributions (Supplementary Figure 4).

Notably, the proportion of African ancestry was strongly increased by means of GMDB micro-publications which account for 40% of the individuals with African ancestry (Figure 4d). In terms of specific sub-ancestries (Figure 4e), more than 80% of cases with sub-Saharan ancestry and over 20% of cases with North African, Native American, and Latin American ancestries were obtained through GMDB micro-publications.

### Performance disparities in underrepresented populations

We analyzed the performance of GestaltMatcher on the test set of 882 images of 275 disorders with different ancestries that have not been used for the training of GestaltMatcher. Performance is measured as a top-k accuracy (as described in Methods). We report the top-1 to top-30 accuracies in Table 1. When considering top-1 accuracy, the ‘Others’ group demonstrated the highest performance at 73.91%, followed by the African group at 62.07%, the Asian group at 53.54%, and the European group at 55.45%. The African group achieved the highest top-5 accuracy (82.76%), the Asian group attained the highest top-10 accuracy (85.04%), while the European group only achieved 75.14% and 82.60% for top-5 and top-10 accuracies, respectively. However, the European group contains more than 50% of the testing images (523 out of 882), covering many more disorders than the other ancestry groups. That includes ultra-rare disorders known to achieve lower performances^19^.

**Table 1.** Performance of GestaltMatcher on different categories of sex, ancestry, and age. The top-1, top-5, top-10, and top-30 accuracy are reported. For the top-1 to top-30 columns, the best performance in each category is boldfaced. In the ancestry category, the sampling influences European and other ancestry groups’ performance due to the significant difference in the test image size. They may evaluate the different sets of disorders. We, therefore, presented the performance of the overlapped disorders in Table 2. In the age category, the notation [x, y) represents a half-open interval, which includes the starting point x but excludes the endpoint y. For example, [0, 1) years range from birth but do not include one year old.

| Category |  | Test images | Top-1 | Top-5 | Top-10 | Top-30 |
| --- | --- | --- | --- | --- | --- | --- |
| Overall |  | 882 | 56.58% | 76.08% | 82.61% | 90.36% |
| Ancestry | African | 29 | 62.07% | **82.76%** | 82.76% | 86.21% |
|  | Asian | 127 | 53.54% | 78.74% | **85.04%** | 89.76% |
|  | European | 523 | 55.45% | 75.14% | 82.60% | 90.25% |
|  | Others | 69 | **73.91%** | 81.16% | 81.16% | **92.75%** |
|  | Unknown | 134 | 53.73% | 73.13% | 81.34% | 91.04% |
| Sex | Male | 419 | 55.37% | 74.22% | 80.67% | 88.78% |
|  | Female | 393 | 55.98% | 75.83% | 83.21% | 91.09% |
|  | Unknown | 70 | **67.14%** | **88.57%** | **91.43%** | **95.71%** |
| Age | $[0, 1)$ years | 53 | 52.83% | 71.70% | 79.25% | 90.57% |
| | $[1, 5)$ years | 137 | 56.20% | 75.91% | 81.02% | 90.51% |
| | $[5, 10)$ years | 115 | 57.39% | **83.48%** | **86.09%** | 90.43% |
| | $[10, \infty)$ years | 165 | **58.18%** | 71.51% | 77.58% | 85.45% |
|  | Unknown | 412 | 56.31% | 76.46% | 84.71% | **92.23%** |

To fairly compare the European group to another non-European ancestry, we only looked at the disorders that were present in both ancestry groups. In Table 2, when comparing the African and European groups on the six overlapping disorders, the European group outperformed the African group by achieving +16.96% top-1 accuracy and +11.17% top-10 accuracy. The European group also exhibited higher accuracies compared to the Asian group, with a top-1 accuracy of +6.92% and a top-10 accuracy of +4.15%. However, the European and ‘Others’ groups achieved relatively comparable results. The ‘Others’ group had a higher top-1 accuracy, while the European group performed better on the top-10 accuracy.

**Table 2.** Performance comparison between European and other ancestry groups on the overlapping disorders. This table is an extension of the ancestry section in Table 1, taking the overlapped disorders between European and other ancestry groups. Each category compares European and non-European ancestry groups’ performance on the same set of disorders. The number of overlapped disorders is reported in the ‘Disorders’ column. In comparing African and European groups, six disorders exist in the test sets of both ancestry groups. The top-1, top-5, top-10, and top-30 accuracy are reported. For the top-1 to top-30 columns, the best performance in each category is boldfaced.

| Category | Disorders |  | Test images | Top-1 | Top-5 | Top-10 | Top-30 |
| --- | --- | --- | --- | --- | --- | --- | --- |
| [African, European] | 6 | African | 14 | 64.29% | 85.71% | 85.71% | 92.86% |
|  |  | European | 32 | **81.25%** | **96.88%** | **96.88%** | **100.00%** |
| [Asian, European] | 36 | Asian | 83 | 57.83% | 79.52% | 84.34% | 87.95% |
|  |  | European | 139 | **64.75%** | **82.73%** | **88.49%** | **91.37%** |
| [Others, European] | 20 | Others | 53 | **81.13%** | **90.57%** | 90.57% | **100.00%** |
|  |  | European | 115 | 69.56% | 84.35% | **93.91%** | 96.52% |
| [Unknown, European] | 32 | Unknown | 77 | 59.74% | **81.81%** | 88.31% | **96.10%** |
|  |  | European | 170 | **62.35%** | 81.18% | **89.41%** | 94.12% |

We further reported the performance of sex and age groups in Table 1. The distribution of testing images was relatively balanced across different groups, and no significant performance gap was observed between males and females. However, the under-one-year-old group exhibited the lowest performance, while the five-to ten-year-old group demonstrated notably higher top-5 and top-10 accuracies.

### Diverse ancestry data enhance prediction accuracy for underrepresented populations

To investigate the impact of incorporating ancestry-diverse data on the overall performance of GestaltMatcher across ancestries, we designed two sets of ancestry analysis experiments. First, we investigated the expansion of the training set of GestaltMatcher (as described in Methods), including either European only (EU + EU*) or European and non-European (EU + non-EU) patients. We measured a top-1 accuracy averaged over all ancestral groups of 49.65% for the European only training set (EU + EU*) and 66.90% for the diverse training set (EU + non-EU) (Figure 5a). Similarly, top-5 accuracy of the European training set was 69.95%, and when we trained on the diverse set, the top-5 accuracy increased to 81.24%. Notably, the evaluation performance on images of patients with European ancestry showed only a marginal performance dropdown. Specifically, the top-1 accuracy decreased by 3.82% and the top-5 accuracy by 3.61% when the dataset was augmented with 50% more non-European images. Meanwhile, the top-1 and top-5 performance increased notably for almost every other ancestral group. Figure 5a and Table 3 show further per-ancestry performances.

**Table 3.** Training accuracy with EU + non-EU and EU + EU* datasets. Within the European training row, numbers annotated with * in brackets indicate the training images from EU + EU. Higher top-1 and top-5 accuracies between EU + EU* and EU + non-EU training are denoted in bold.

|  | Number of images |  | Performance EU + non-EU |  | Performance EU + EU* |  |
| --- | --- | --- | --- | --- | --- | --- |
|  | Training | Testing | Top-1 | Top-5 | Top-1 | Top-5 |
| European | (4706.2 ± 24.4)*<br>3139.2 ± 15.1 | 444.6 ± 22.2 | 52.35 ± 2.30% | 72.05 ± 2.66% | **56.17 ± 2.27%** | **75.66 ± 2.70%** |
| East Asian | 283.2 ± 5.0 | 31 ± 6.2 | **55.78 ± 10.25%** | **74.56 ± 5.90%** | 37.77 ± 5.45% | 60.13 ± 5.77% |
| Latin/Hispanic | 257.8 ± 7.0 | 28.4 ± 4.7 | **68.86 ± 8.92%** | **82.56 ± 7.77%** | 66.16 ± 7.89% | 80.51 ± 6.58% |
| Middle-East/<br>West Asian | 211.2 ± 6.8 | 30 ± 5.8 | **46.76 ± 7.01%** | **67.59 ± 7.52%** | 36.10 ± 6.81% | 59.67 ± 3.68% |
| South Asian | 200.2 ± 5.4 | 18.8 ± 2.7 | **72.15 ± 12.24%** | **87.32 ± 10.04%** | 53.70 ± 14.86% | 66.13 ± 13.40% |
| Asian Others | 170.6 ± 2.4 | 16.4 ± 4.1 | **64.66 ± 10.93%** | **80.06 ± 13.87%** | 41.64 ± 18.51% | 66.84 ± 11.87% |
| Sub-Saharan | 119 ± 2.7 | 18.2 ± 3.4 | **54.23 ± 7.32%** | **75.49 ± 15.27%** | 28.91 ± 11.18% | 46.55 ± 11.71% |
| North African | 64.8 ± 3.2 | 7.4 ± 1.6 | 79.64 ± 20.19% | 86.64 ± 14.62% | 42.71 ± 19.34% | 71.64 ± 18.38% |
| Native American | 63.2 ± 6.8 | 14.8 ± 1.6 | 84.55 ± 12.77% | 99.09 ± 1.82% | 83.94 ± 11.18% | 94.65 ± 8.62% |
| African Others | 53.2 ± 2.0 | 6.2 ± 2.2 | 72.78 ± 18.29% | 85.00 ± 13.33% | 55.56 ± 17.57% | 73.33 ± 22.61% |
| South-East Asian | 51.4 ± 2.0 | 5.4 ± 1.3 | 72.12 ± 13.36% | 78.81 ± 24.61% | 24.40 ± 6.76% | 46.83 ± 17.97% |
| Others | 54.6 ± 2.3 | 6 ± 2.9 | 68.71 ± 23.67% | 79.43 ± 22.38% | 59.00 ± 22.35% | 71.57 ± 21.09% |
| African American | 38.4 ± 4.3 | 3.8 ± 2.6 | 77.08 ± 18.04% | 87.50 ± 21.65% | 59.38 ± 24.00% | 69.79 ± 18.49% |
| Overall | 4706.8 ± 26.7 | 631 ± 23.8 | 66.90% | 81.24% | 49.65% | 67.95% |

**Figure 5.**
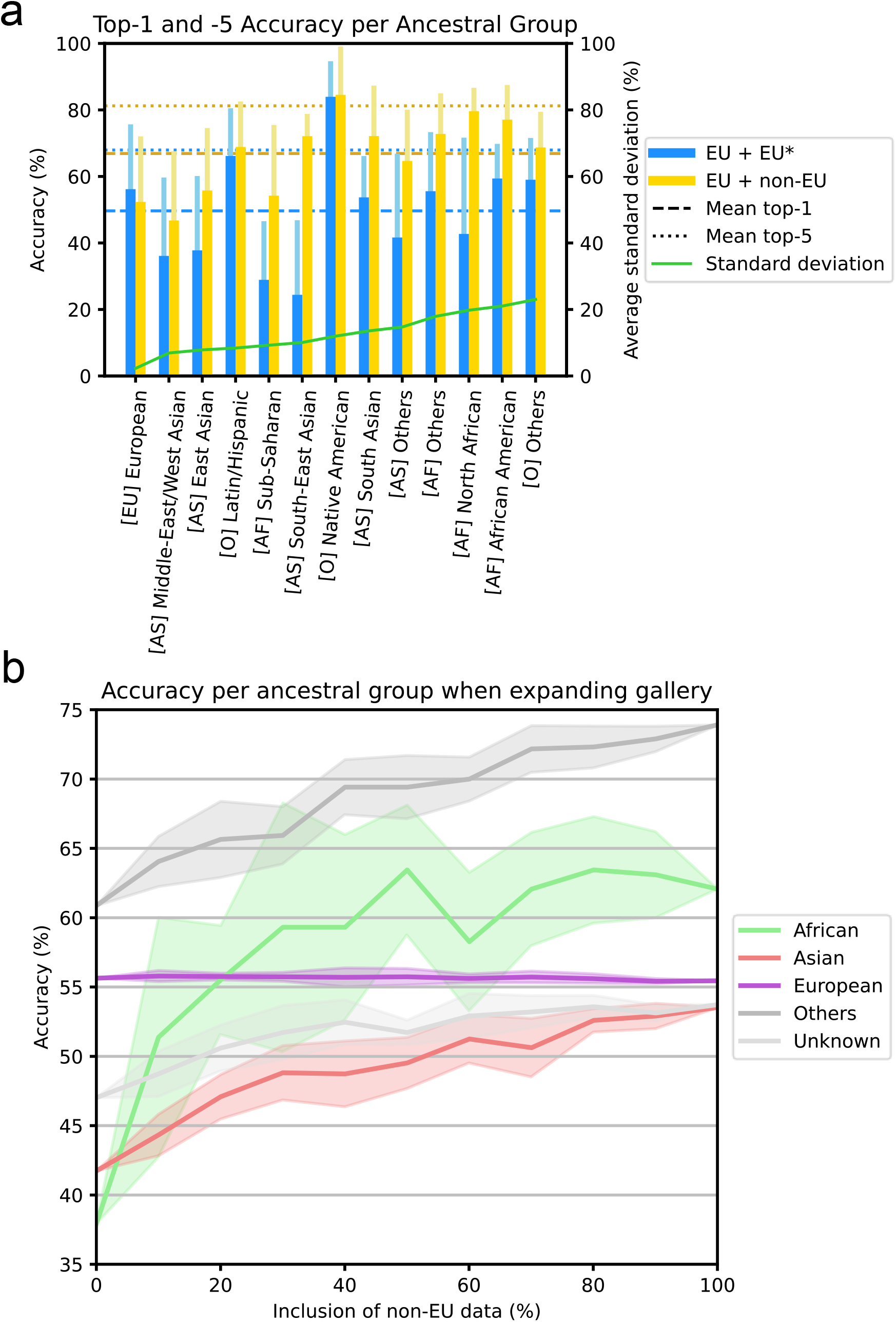
Performance of ancestry analysis. **a)** Top-1 and top-5 accuracy of GestaltMatchers’ disorder classification accuracy per ancestral group. Top-1 and top-5 accuracy of the models’ disorder classification accuracy per ancestral group, where (blue) belongs to the EU only subset, and (yellow) belongs to the diverse subset. Each wide, darker bar and each light, thinner bar indicate the top-1 and top-5 accuracy per ancestral group, respectively. The horizontal dashed lines and dotted lines indicate the top-1 and top-5 overall accuracy averaged over all ancestral groups, respectively. The order of the ancestry group in the x-axis is ranked according to standard deviation between top-1 accuracies of the 5-fold experiment. **b)** Top-1 accuracy of GestaltMatcher when including different proportion of non-European patients in the gallery. The x-axis is the proportion of non-European data included in the gallery. The y-axis is the top-1 accuracy. The colored region along the line indicates the standard deviation.

The training of GestaltMatcher results in a clinical face phenotype space that can be populated by additional cases, which we refer to as the gallery set (as described in Methods). We next investigated the influence of expanding the gallery with ancestry-diverse data by gradually raising the proportion of included non-European data from 10% to 100%. Figure 5b shows that the top-1 accuracy of the non-European groups was clearly increased when we added more non-European data in the gallery. However, the top-1 accuracy of the European group did not change even when we added 100% of the non-European data into the gallery.

### GMDB-FAIR dataset drives the advancement of NGP technology

GMDB-FAIR dataset is the first dataset that can be shared with the research community to train and benchmark their NGP approaches. After the first publication of the GestaltMatcher approach in 2022, for which we initially started the collection of our FAIR data, many researchers have utilized GMDB-FAIR to develop different NGP approaches. Hustinx et al.^19^, Sumer et al.^35^, and Campbell et al.^36^ improved the prediction accuracy of their models significantly by utilizing different loss functions, network architectures, and data augmentation. Recently, Wu et al. proposed combining a large language model with facial image analysis to streamline the rare disorder diagnosis^37^. Furthermore, running facial analysis with an on-premise solution is possible using the FAIR data set to further prioritize genomic variants^38^.

Moreover, the GMDB-FAIR dataset can be taken as a validatable control cohort to facilitate the delineation of the facial phenotype of disorders. GestaltMatcher can detect clusters and assess whether, for example, cases with an identical variant or pathogenic variants in the same gene share a similar facial phenotype. For example, Ebstein et al. showed that facial dysmorphism was heterogeneous among the entire *PSMC3* patient cohort, but facial similarities were found in patients sharing the same pathogenic variants^39^. To date, 15 publications have analyzed the facial phenotype of the cohort with the GMDB-FAIR dataset and GestaltMatcher^39–53^. All results can be reproduced in the research platform of GMDB, which we introduce in the Methods section (Figure 2c, Figure 3c and Supplementary Note).

## Discussion

GMDB is a modern, searchable reference and publication medium encompassing diverse populations that is designed for both clinicians and computer scientists engaged in NGP development. The ultimate goal of this study is to drive research in rare genetic disorders to understand the phenotypic variability among ancestries systematically and improve support for underrepresented populations.

GMDB stands out as the sole database compliant with FAIR principles, distinguished by its extensive collection of facial images covering diverse populations. This was mainly possible through the contributions and crowd-sourced annotations by our global collaborators. To increase motivation for data submission in the future, every case in the database has the potential to become a citable micro-publication with a Digital Object Identifier (DOI)^54^. Furthermore, future micro-publications could be indexed in reputable scientific indexing services, such as PubMed, as is the case for some existing micro-publication communication platforms^55^. Active patient involvement and the ability to access, upload and delete their data enhance patient autonomy and facilitate the acquisition of longitudinal patient data, further enriching GMDB’s repository of facial images. Similar to other natural history study data, the longitudinal image and associated phenotypic meta data add significant value to the understanding of disease progression in patients with facial dysmorphism^56^. Moreover, micro-publication encourages the recruitment of patients from underrepresented populations. For example, more than 40% of all images obtained for Africans had been previously unpublished. These micro-publications from unpublished images of patients with underrepresented ancestries underscored the importance of GMDB since they cannot be found in any medical journals.

The diverse ancestry data in GMDB further enabled us to investigate the GestaltMatcher performance differences among ancestral groups systematically. In Table 2, the performance disparities in the Asian and African groups were observed when compared to the European group. The “Others” group showed a comparable or even higher performance than the European group. The reason could be that Latin Americans in the ‘Others’ group show relatively similar facial phenotypes to the Europeans.

Our findings indicate that increasing the ancestral diversity in FAIR databases will particularly benefit populations currently regarded as underprivileged. We investigated how the top-1 and top-5 accuracies for the different ancestries changed when equally sized groups of European or non-European patients were added to the training set. Overall, the top-5 accuracy for non-European ancestral groups increased significantly when the training set was expanded with non-Europeans (+11.29%). When the training data were extended from only Europeans to Europeans and non-Europeans, only a marginal change in the performance of the European group was observed. Including more non-European patients in the gallery can also improve non-European groups’ performances dramatically while European performance remains roughly the same (Figure 5b). The results indicate that recruiting non-European patients to support the underrepresented populations is more effective than recruiting more European patients, which often leads to models’ extreme bias toward European ancestry.

The GMDB-FAIR dataset offers a transparent AI training set, which is crucial for the NGP development because all FAIR data are available to the clinical and scientific community. This transparency, combined with the increased representativeness of the training set, helps minimise the risk of algorithmic bias, which is key for ensuring respect for the fundamental right to non-discrimination^57^. The high quality of the GMDB data allows researchers to train, validate, and test AI in a manner that aligns with the expectation in the EU AI Act and the EU Medical Device Regulation^58^. Finally, the controlled access and consent options as described in the Methods section not only ensures respect for the fundamental right to protection of personal data^57^ and EU General Data Protection Regulation (GDPR)^59^ compliance, but it also enabled the creation of a more diverse, representative, and larger data set as people are more comfortable with sharing health and genetic data, including images, under controlled conditions and responsible data governance than in open access publications and repositories. By this, the GMDB-FAIR dataset falls in line with other large public datasets, such as ImageNet^60^ for object classification or Labeled Faces in the Wild (LFW)^61^ for face verification, which have been fundamental for deep-learning technology driving computer vision over the last decade. GMDB-FAIR has been used to develop many NGP approaches^19,35–37^ for predicting rare disorders after the first usage in GestaltMatcher in 2022. Moreover, GMDB-FAIR data can be used in the research platform (Supplementary Note) to validate the results shown in the published works^39–53^ that provides transparency to the researcher using GestaltMatcher and the probability to extend the existing research with the user’s additional data.

Due to variability in facial phenotypes secondary to ancestry, diverse reference image databases are crucial in order to enable clinicians to learn about the phenotypic variability in facial dysmorphism within a given disorder. While efforts have been made to create an atlas of human malformations that addresses the issue of ancestral diversity, this remains limited to only a few disorders^20^. With GMDB-FAIR, we created a large-scale dataset that can be searched for disorders or genes of interest in the GMDB gallery view (Figure 2c, Figure 3b), which provides clinicians with a comprehensive selection of patient images from different ancestries at a glance, thereby eliminating the need for extensive literature searches. In addition, it facilitates facial phenotype comparisons within a given disorder among different ancestries (Supplementary Note). GMDB also represents a valuable teaching tool for training students and residents to recognize disorders based on facial features.

To conclude, GMDB is a medical imaging database for rare disorders that encompasses diverse populations. The FAIR data will serve as reference material for clinicians that facilitates learning about facial dysmorphism across ancestries, and as a transparent training and benchmarking dataset for advancing the NGP approach. While we show improved performance for the underrepresented populations, it is important to point out that the performance is far from the optimum that can be achieved by collecting more diverse data. We envision that the gap between the European ancestral group and the underrepresented ancestries can be mitigated by micro-publications in the future, and this will result in substantially improved support for underrepresented populations.

## Methods

### Implementation of the online GMDB platform

The online platform was built using Ruby on Rails in order to allow users to input images and other patient data. A database was set up using MySQL to store the patient data. GMDB is hosted physically in the University Hospital of Bonn and is maintained by Arbeitsgemeinschaft für Gen-Diagnostik e.V. (AGD), which is a non-profit organization for genomic research. The service is funded by membership fees of the AGD and donations from the Eva-Luise und Horst Köhler Foundation and the Wirtgen Foundation.

### Image data and meta data stored in GMDB

An entry in GMDB consists of a medical image such as a portrait, X-ray, or fundoscopy and machine-readable meta information containing: 1) demographic data (including sex, age, and ancestry); 2) the molecularly confirmed diagnosis (OMIM index^62^); 3) the disease-causing mutation reported in Human Genome Variation Society format^63^ (HGVS) or International System for Human Cytogenomic Nomenclature^64^ (ISCN) with test method and zygosity; and 4) the clinical feature encoded in HPO terminology^34^ (Figure 2b). When submitting data, clinicians are also asked to state their expert opinion concerning the distinctiveness of a phenotype: They are asked to score whether the medical imaging data was supportive (1), important (2), or key (3) in establishing the clinical diagnosis. Computer scientists can use this information to interpret the performance of their AI^15^.

### Digital consent form and patient-centered data upload

To facilitate faster retrospective patient recruitment, a digital consent form has been implemented, which allows patients to select conditions for storing their data within the database and enables the provision of their signature online. To address the specific requests of patients, this feature was further developed in close collaboration with patient support groups, e.g., the German Smith-Magenis Syndrome patient organization Sirius e.V. Patients can access their own cases and provide or withdraw their consent online. They can also upload images themselves, which greatly simplifies the curation process for longitudinal image data and other prospective data. The fact that documents such as letters from clinicians or laboratory results can also be uploaded, while only being visible to the responsible clinician, makes it possible to obtain molecular and phenotype information on patients recruited retrospectively from patient support groups. This digital consent is developed in such a way that it could also, in principle, be used as a dynamic consent model in the future^65^. The consent form is available in German and English, and other languages will be incorporated in the near future. Please find them in Supplementary Note (Digital consent, and Supplementary Figures 5 and 6) for more details.

### Data curation

The curated data can be broadly categorized as retrospective and prospective. Retrospective refers primarily to data collected from the literature or from similar projects with global consent for data sharing (e.g., Minerva&Me^66^). For cases curated from the literature, the DOI and PubMed ID as well as the contact details of the corresponding author were collected in order to clarify whether reuse is possible while respecting intellectual property rights. Following the provision of written informed consent, our collaboration partners, clinicians from around the world (Figure 1a and the co-authors), also recruited patients with an established diagnosis from within their clinical practice or from patient support groups. Prospective curation refers to the collection of further images or metadata over time. This can be done by the attending clinician after subsequent consultations, or by the patients themselves.

The curation process can be broadly subdivided into three phases. First, medical students in their final year annotated cases from the literature, mainly searched PubMed and Google Scholar for publications with images of patients with facial dysmorphism and monogenic molecular diagnosis.

Second, solved patients were recruited from patient support groups. Included patients were allowed to upload and delete images and findings autonomously and access their data at any time. To develop a patient-centered, user-friendly platform and strengthen patient autonomy, feedback was obtained from the recruited patients during this phase in order to determine whether any adjustments to the process were required.

In the third phase, the database was expanded via international collaborations with clinicians from different continents. Initially, this focused on patients who had already been solved but had not yet been published in order to improve the AI’s performance. However, as we progressed, more clinicians shared their unsolved cases with the scientific community. GMDB then started focusing on facial portraits of patients with rare monogenic diseases, and is now dominated by, but not limited to, such cases. Later in the curation process, we also annotated cytogenetic disorders with facial dysmorphism. In addition to these clinicians, the medical students continued to annotate data from the literature.

### Digital Object Identifier assignment

After data submission, the respective case is immediately published on the website. Subsequently, the author has the option of generating a DOI in order to create a citable micro-publication^54^. To do this, clinicians must, after uploading the required data and metadata, enter their own personal identifier (e.g.,ORCID), specify all other scientists or clinicians involved in this case, and provide a title and an abstract. To ensure the credibility and reliability of the published data, this process will adhere to a rigorous review similar to that described by Raciti et al.^55^. The DOIs are created and managed by the University and State Library of Bonn using the DataCite Application Programming Interface (API) (https://datacite.org).

Additionally, a dedicated landing page will be created for each case, according to the specifications of the DataCite metadata schema (Supplementary Figure 2). The landing page is accessible via the generated DOI, even for individuals without access to GMDB or those who are not logged in. The landing page contains the full citation with the DOI as a link, the abstract, and a description of the case data. No phenotypic information, HPO terms, or images are available. However, the landing page indicates how many images the micropublication contains.

### Main components of the GMDB online platform

The GMDB consists of three main components that can in principle be utilized by registered users (Figure 2c). 1) Search: Clinicians can use the Gallery view to search the GMDB for disorders or genes of interest and get all patients matching this search criterion displayed in the database at a glance. 2) Analyze: Clinicians and scientists can use the GMDB-FAIR data to perform similarity comparisons of cohorts with GestaltMatcher within the research platform of GMDB. 3) Train: The GMDB-FAIR dataset that can be used by external researchers to train NGP tools. More detailed information on these features can be found in the Supplement Note.

### GMDB datasets

All analyses performed in this paper are based on GMDB-FAIR data (v1.1.0). But actually, the GMDB consists of the GMDB-FAIR dataset and the GMDB-private set (Supplementary Note and Supplementary Figures 7 and 8). We introduced this distinction because it is known that patient consent to data sharing is higher when not shared with a broad mass, but only for a specific study^67^. However, many patients agree to controlled access for the general scientific community to advance research^67^. For this reason, patients can decide whether they want to be part of only the GMDB-private set for AI training or agree to be part of the FAIR data set.

The website displays the statistics to the public, showing how many patients are in the database and how many disorders and disease genes have been curated. When the user has the link to a specific case in the GMDB (e.g., from a publication in which the original image may not be branched, but a link to the case is given in the GMDB), if the user is not logged in, the landing page for the case will show how many images and metadata are available for the case. Only sex and ancestry, as well as the disease gene, are given. If it is a case report published with a DOI in the GMDB, the corresponding title and abstract of the case can also be viewed. The remaining data can only be viewed after logging in. To visualize the images, the user has to log in to the platform.

### GMDB-FAIR data set

The FAIR data set (Supplementary Figure 7b) is accessible to the scientific community. Data comes from publications and from clinicians or patients themselves. However, the case is accessible in the Gallery view for all registered users of the GMDB, and the data sheet with all relevant data and metadata can be viewed. It is also available to all users of the GMDB to perform similarity comparisons of cohorts in the research platform (Supplementary Note). The data is used for the GestaltMatcher training and test set but can also be made available to other scientists to train and test their AI after they have applied to us with an Institutional Review Board (IRB)-approved study and proposal.

### Data Governance and Ethical, Legal and Social Implications of GMDB

Ethical approval for the GMDB was granted by the IRB of the University of Bonn, and all patients have given informed written consent to participate. During the GestaltMatcher consent procedure, patients can also indicate whether they agree to the use of the images in presentations, teaching activities, or in publications in other journals. This differentiation from other journals is important since patients/parents show less willingness to consent to publication in open-access journals than to publication in access-controlled databases that are not publicly accessible^67^. The patient shown in Figure 3 fully consented to publication of his image data.

The GMDB has four different levels of data access (Supplementary Figure 8): 1) The public data, which includes a summary of the GMDB statistics on the website and a landing page for case reports with DOI (Supplementary Figure 2), requires no login and is openly accessible. 2) The FAIR data, which can be viewed with a GMDB user account, and in principle, downloaded by external AI researchers. 3) The restricted data, which is not accessible to GMDB users and external AI researchers and can only be used to train the GestaltMatcher AI. 4) Patient-shared data: Patients can only view their own case and upload data if they are invited to do so by the attending clinician.

External scientist in the field of AI can apply to download of GMDB-FAIR data for the development of NGP approaches. Prerequisites for this are IRB approval and submission of a proposal to. In addition, external scientists must sign and adhere to the GDPR. The Advisory Board will conduct a thorough review of all applications. If the majority of the members of the Board approve the application, access (under the extent permissable by law) will be granted to applicants within two to three weeks.

### Advisory Board

Advisory Board comprises the following co-authors: Benjamin D. Solomon, Koen Devriendt, Shahida Moosa, Christian Netzer, Martin Mücke, Christian Schaaf, Alain Verloes, Christoffer Nellåker, Markus M. Nöthen, Gholson J. Lyon, Aleksandra Jezela-Stanek, and Karen W. Gripp.

### HPO-defined symptom groups

In one of our previous works^68^, twelve distinct and non-overlapping categories of HPO terms were defined by clinical experts (“HPO defined symptom groups”). All GMDB cases for which HPO terms were annotated were then assigned to each of those groups, if at least one of the HPO terms in this group was annotated; i.e., each GMDB case can be assigned to several HPO-defined symptom groups. For each case, the most pronounced HPO-defined symptom group was defined as the single group comprising the largest number of the case’s annotated HPO terms. The HPO-defined symptom group “Others” was only assigned as the leading HPO-defined symptom group if no other HPO-defined symptom group was present for the case.

Phenotypic similarity between cases was calculated using the R-package ontologySimilarity (version 2.5). Pairwise similarities were calculated for the combined data set of GMDB cases with HPO terms (n=4,474), the TRANSLATE-NAMSE exome sequencing data set (n=1,577), and data on known diseases and their clinical features downloaded from the HPO website (n=7,765, https://hpo.jax.org/app/download/annotation, file: genes_to_phenotype.txt, downloaded on 10 April 2021). The resulting distance matrix was projected in a four-dimensional space via Uniform Manifold Approximation and Projection (UMAP). The first two dimensions were plotted using ggplot2 (version 3.4.4). To analyze which HPO-defined symptom groups occur jointly, the proportion of patients assigned to the first group that were also assigned to the second group was assessed. All analyses were conducted in R (version 4.3.2).

### GestaltMatcher Algorithm

GestaltMatcher^15^ is the extension of the DeepGestalt approach^17^. DeepGestalt is a deep learning-based NGP tool using frontal face photos to classify up to 216 syndromes it has seen during training. However, it needed a lot of training data to achieve a reasonable performance on these syndromes. That also meant it could not classify unseen syndromes during training (ultra-rare syndromes). This led to the development of GestaltMatcher, which uses a clustering approach. As such, if at least one image of the sought-after syndrome is in the gallery set, a test image can be matched to/clustered with that image using some similarity metric. Later, this approach was further enhanced by Hustinx et al.^19^, using a more recent architecture (iResNet) and training loss (ArcFace Loss), as well as test-time augmentation and a model ensemble to improve robustness. That is also the approach we used for our experiments. Thus, for fine-tuning, we utilized the Adam optimizer, cross-entropy loss, and class weighting to deal with the imbalance in data availability between disorders. In this study, we used 7,787 images representing 275 disorders as the training set and a validation set of 1,007 images during the model training. We then tested the model on a test set consisting of 882 images.

The overall idea behind the methodology is to train a classifier on a more frequent subset of the syndromes, achieving a model that generalizes well on those seen syndromes. In practice, the authors of both papers decided to use syndromes with at least seven patients as the training set for this classifier. Thereafter, everything up to the penultimate layer of the classifier is used as an encoder, obtaining feature embeddings of images of interest. These could be images for the gallery set or images for the test set.

The aforementioned gallery set is the set of images (and their feature embeddings) with known syndromes. This can include the syndromes used for training (seen) and syndromes with too few images to train on (unseen). The theory is that similar facial phenotypes form clusters in the feature space, which is spanned by the feature embeddings in 512 dimensions and which we refer to as clinical face phenotype space. The similarity between images and clusters is computed using the cosine distance, where a lower distance implies a higher similarity. Contrary to the approach by Gurovich et al.^17^, this approach can easily increase support for ultra-rare syndromes. The quality and diversity of the gallery set is crucial for this approach to match test images to clusters in the gallery set.

### Performance metric (top-k accuracy)

The applied performance metric was top-k accuracy. Top-1 indicates that the disorder was correctly classified as the first guess, while top-5 indicates the correct class was in the first five guesses. We reported top-k accuracies (k=1, 5, 10, and 30) as the performance readout.

### Ancestry analysis

The genetic ancestry of each individual was documented as precisely as possible using self-reported data. For instance, if an individual was born in Germany and all of the respective grandparents also originated from there, this individual was assigned to Germany (country) and Europe (continent). The same approach was used for all individuals with no self-reported migration history in previous generations. For individuals with mixed ancestry, the respective ancestries were combined. For example, an individual with a father from Gambia and a mother from Eastern Europe was assigned European-African mixed ancestry.

The performance of GestaltMatcher is highly dependent on the training set and the gallery set. To investigate the impact of incorporating diverse ancestry on the performance, we have therefore conducted two sets of experiments for those two components, respectively. First, we analyzed the influence on the models’ performance when including only European versus both European and non-European data into the training set. And second, we analyzed the same performance when iteratively increasing the amount of non-European data into the gallery set.

In the first experiment, a subset of images of European patients (EU) was extended by either the inclusion of a different subset of images of European patients (EU*), or a subset of patients with non-European ancestries (non-EU) (Supplementary Figure 9). Random sampling of these subsets was performed five times. EU consisted of on average 3,139.2 images, and EU* comprised on average 1,567.6 images. First, the model was trained on the EU + EU* set containing on average 4,706.8 images of patients of solely European ancestry. For EU + non-EU, a subset containing on average 1,567.6 images of patients with any non-European ancestry was used, totaling to 4,706.8 images. The experiment design ensured the maintenance of the same distribution of disorders as that found in the training data.

The model was fine-tuned for 50 epochs on subsets EU + EU* and EU + non-EU of GMDB (v1.1.0). All other hyperparameters were left unchanged. It is important to note that the model was not tasked with learning to classify the ancestry, only with learning to classify the disorder.

Post-training, the models’ performances were measured on the same evaluation set, containing images of patients with diverse ancestral backgrounds. This evaluation set consisted of 649 images and was sampled in such a manner that there was no overlap between patients or images in any subset. Top-k accuracy was averaged over each ancestry rather than each image in order to address the imbalance in ancestry frequency. As such, the performance of any infrequent group weighed equally with those of the more frequent groups.

In the second set of experiments, we trained the models of Hustinx et al.^19^ using the GMDB-FAIR training set, including different proportions of non-EU data for the gallery set. We compared the performance of the syndromes our models have seen during training. For completeness, Table 1 shows the top-k accuracy (over all images) for different categories (sex, ancestry, and age range) using the entire gallery set consisting of 8,794 images (100% EU [4911] + 100% non-EU [3883]). For the experiments, we computed the performance when including different proportions of non-EU data, extending the gallery set by +10% per iteration. This experiment was repeated tenfold, randomly sampling patients with different ancestries and all their photos for the gallery set. As such, at 0%, we include only data from EU patients in the gallery set, and at 100%, we include all patient data for the relevant syndromes.

We further computed the performance on syndromes that occur in both the European-group and each non-European group to more accurately reflect the performance differences, avoiding the imbalance between offered support for each ancestral group.

## Data Availability

All data produced are available online at https://db.gestaltmatcher.org.

## Data and code availability

GMDB-FAIR can be downloaded in GMDB after the application is approved by the advisory board. Please find more details in the Data Governance and ELSI section. Code is available in the GitHub repository (github.com/igsb/GestaltMatcher-Arc/tree/gmdb).

## Acknowledgments

This research was supported in part by the Intramural Research Program of the National Human Genome Research Institute, National Institutes of Health, the United States of America. Tzung-Chien Hsieh, Peter M. Krawitz and Annabelle Arlt are partner of the European Joint Programme on Rare Diseases (EJP RD) for the project ANR-22-RAR4-0001-01 (UPS36NDDiag). Sofia Douzgou Houge was supported by the Norwegian National Advisory Unit on Rare Disorders (grant number #43066). Tahsin Stefan Barakat was supported by the Netherlands Organisation for Scientific Research (ZonMw Vidi, grant 09150172110002). Heidi Beate Bentzen is supported by EU grant 101071203 and Research Council of Norway grants 322672 and 324278. Nina-Maria Wilpert was supported by the DFG Research Unit 2841 “Beyond the Exome” and is a participant in the BIH Charité Junior Clinician Scientist Program funded by the Charité - Universitätsmedizin Berlin, the Berlin Institute of Health at Charité (BIH), the Alliance4Rare, and the Berliner Sparkassenstiftung Medizin. Cristina Dias was supported by the Wellcome Trust [grant number 209568/Z/17/Z]. The authors thank the Asia Pacific Society of Human Genetics, the Wirtgen Foundation, the Eva Luise und Horst Köhler Foundation, Kabuki Syndrome Foundation, Kleefstra support group, German Smith-Magenis Syndrome patient organization Sirius e.V. and the Focus Foundation for their support.

## Notes

### Competing Interest Statement

The authors have declared no competing interest.

### Funding Statement

This study did not receive any funding.

### Author Declarations

Ethics committee/IRB of University Hospital of Bonn gave ethical approval for this work

### Summary of Updates

Update the main manuscript to the submission version.

## References

1. Hart, T. C. & Hart, P. S. Genetic studies of craniofacial anomalies: clinical implications and applications. Orthod. Craniofac. Res. 12, 212–220 (2009).

2. Lesmann, H., Klinkhammer, H. & Dr. med. Dipl. Phys. Peter M. Krawitz. The future role of facial image analysis in ACMG classification guidelines. Med. Genet. 35, 115–121 (2023).

3. Tekendo-Ngongang, C. et al. Rubinstein-Taybi syndrome in diverse populations. Am. J. Med. Genet. A 182, 2939–2950 (2020).

4. Kruszka, P., Tekendo-Ngongang, C. & Muenke, M. Diversity and dysmorphology. Curr. Opin. Pediatr. 31, 702–707 (2019).

5. Hadj-Rabia, S. et al. Automatic recognition of the XLHED phenotype from facial images. Am. J. Med. Genet. A 173, 2408–2414 (2017).

6. Martínez-Abadías, N. et al. Facial biomarkers detect gender-specific traits for bipolar disorder. FASEB J. 35, (2021).

7. Fang, F., Clapham, P. J. & Chung, K. C. A systematic review of interethnic variability in facial dimensions. Plast. Reconstr. Surg. 127, 874–881 (2011).

8. Vorravanpreecha, N., Lertboonnum, T., Rodjanadit, R., Sriplienchan, P. & Rojnueangnit, K. Studying Down syndrome recognition probabilities in Thai children with de-identified computer-aided facial analysis. Am. J. Med. Genet. A 176, 1935–1940 (2018).

9. Kruszka, P. et al. Down syndrome in diverse populations. Am. J. Med. Genet. A 173, 42–53 (2017).

10. Porras, A. R., Summar, M. & Linguraru, M. G. Objective differential diagnosis of Noonan and Williams-Beuren syndromes in diverse populations using quantitative facial phenotyping. Mol Genet Genomic Med 9, e1636 (2021).

11. Lumaka, A. et al. Facial dysmorphism is influenced by ethnic background of the patient and of the evaluator. Clin. Genet. 92, 166–171 (2017).

12. Burchard, E. G. et al. The importance of race and ethnic background in biomedical research and clinical practice. N. Engl. J. Med. 348, 1170–1175 (2003).

13. Martínez-Abadías, N. et al. Phenotypic evolution of human craniofacial morphology after admixture: a geometric morphometrics approach. Am. J. Phys. Anthropol. 129, 387–398 (2006).

14. Fatumo, S. et al. A roadmap to increase diversity in genomic studies. Nat. Med. 28, 243–250 (2022).

15. Hsieh, T.-C. et al. GestaltMatcher facilitates rare disease matching using facial phenotype descriptors. Nat. Genet. 54, 349–357 (2022).

16. Dudding-Byth, T. et al. Computer face-matching technology using two-dimensional photographs accurately matches the facial gestalt of unrelated individuals with the same syndromic form of intellectual disability. BMC Biotechnol. 17, 90 (2017).

17. Gurovich, Y. et al. Identifying facial phenotypes of genetic disorders using deep learning. Nat. Med. 25, 60–64 (2019).

18. Porras, A. R., Rosenbaum, K., Tor-Diez, C., Summar, M. & Linguraru, M. G. Development and evaluation of a machine learning-based point-of-care screening tool for genetic syndromes in children: a multinational retrospective study. Lancet Digit Health (2021) doi:10.1016/S2589-7500(21)00137-0.

19. Hustinx, A. et al. Improving Deep Facial Phenotyping for Ultra-rare Disorder Verification Using Model Ensembles. in 2023 IEEE/CVF Winter Conference on Applications of Computer Vision (WACV) 5007–5017 (IEEE, 2023).

20. Muenke, M., Adeyemo, A. & Kruszka, P. An electronic atlas of human malformation syndromes in diverse populations. Genet. Med. 18, 1085–1087 (2016).

21. Mishima, H. et al. Evaluation of Face2Gene using facial images of patients with congenital dysmorphic syndromes recruited in Japan. J. Hum. Genet. 64, 789–794 (2019).

22. Narayanan, D. L. et al. Computer-aided Facial Analysis in Diagnosing Dysmorphic Syndromes in Indian Children. Indian Pediatr. 56, 1017–1019 (2019).

23. Elmas, M. & Gogus, B. Success of Face Analysis Technology in Rare Genetic Diseases Diagnosed by Whole-Exome Sequencing: A Single-Center Experience. Mol. Syndromol. 11, 4–14 (2020).

24. Hennocq, Q. et al. Next generation phenotyping for diagnosis and phenotype-genotype correlations in Kabuki syndrome. Sci. Rep. 14, 2330 (2024).

25. 1000 Genomes Project Consortium et al. A global reference for human genetic variation. Nature 526, 68–74 (2015).

26. Winter, R. M. & Baraitser, M. The London Dysmorphology Database. J. Med. Genet. 24, 509–510 (1987).

27. Murdoch Children’s Research Institute. POSSUMweb. POSSUMweb https://www.possum.net.au/.

28. Patrinos, G. P. Chapter 6 - Incentives for Human Genome Variation Data Sharing. in Human Genome Informatics (eds. Lambert, C. G., Baker, D. J. & Patrinos, G. P.) 109–129 (Academic Press, 2018).

29. Mons, B. et al. The value of data. Nat. Genet. 43, 281–283 (2011).

30. Patrinos, G. P. et al. Microattribution and nanopublication as means to incentivize the placement of human genome variation data into the public domain. Hum. Mutat. 33, 1503–1512 (2012).

31. Giardine, B. et al. Systematic documentation and analysis of human genetic variation in hemoglobinopathies using the microattribution approach. Nat. Genet. 43, 295–301 (2011).

32. Wilkinson, M. D. et al. The FAIR Guiding Principles for scientific data management and stewardship. Sci Data 3, 160018 (2016).

33. Lesmann, H. & Weiland, H. Atypical presentation of a case with Noonan syndrome with multiple lentigines (Version 1). (2024) doi:10.60723/10693.

34. Robinson, P. N. et al. The Human Phenotype Ontology: a tool for annotating and analyzing human hereditary disease. Am. J. Hum. Genet. 83, 610–615 (2008).

35. Sümer, Ö., Hellmann, F., Hustinx, A., Hsieh, T.-C. & Krawitz, P. Few-Shot Meta-Learning for Recognizing Facial Phenotypes of Genetic Disorders. in Caring is Sharing – Exploiting the Value in Data for Health and Innovation 932–936 (IOS Press, 2023).

36. Campbell, J., Dawson, M., Zisserman, A., Xie, W. & Nellåker, C. Deep Facial Phenotyping with Mixup Augmentation. in Medical Image Understanding and Analysis 133–144 (Springer Nature Switzerland, 2024).

37. Wu, D. et al. Multimodal Machine Learning Combining Facial Images and Clinical Texts Improves Diagnosis of Rare Genetic Diseases. arXiv [q-bio.QM] (2023).

38. Hsieh, T.-C., Lesmann, H. & Krawitz, P. M. Facilitating the Molecular Diagnosis of Rare Genetic Disorders Through Facial Phenotypic Scores. Curr Protoc 3, e906 (2023).

39. Ebstein, F. et al. PSMC3 proteasome subunit variants are associated with neurodevelopmental delay and type I interferon production. Sci. Transl. Med. 15, eabo3189 (2023).

40. Asif, M. et al. De novo variants of CSNK2B cause a new intellectual disability-craniodigital syndrome by disrupting the canonical Wnt signaling pathway. HGG Adv 3, 100111 (2022).

41. Kampmeier, A. et al. PHIP-associated Chung-Jansen syndrome: Report of 23 new individuals. Front Cell Dev Biol 10, 1020609 (2022).

42. Lyon, G. J. et al. Expanding the phenotypic spectrum of NAA10-related neurodevelopmental syndrome and NAA15-related neurodevelopmental syndrome. Eur. J. Hum. Genet. 31, 824–833 (2023).

43. Aerden, M. et al. The neurodevelopmental and facial phenotype in individuals with a TRIP12 variant. Eur. J. Hum. Genet. 31, 461–468 (2023).

44. Blackburn, P. R. et al. Loss-of-function variants in CUL3 cause a syndromic neurodevelopmental disorder. medRxiv (2023) doi:10.1101/2023.06.13.23290941.

45. Oppermann, H. et al. CUX1-related neurodevelopmental disorder: deep insights into phenotype-genotype spectrum and underlying pathology. Eur. J. Hum. Genet. 31, 1251–1260 (2023).

46. Blackburn, P. R. et al. Loss-of-function variants inCUL3cause a syndromic neurodevelopmental disorder. medRxiv (2023) doi:10.1101/2023.06.13.23290941.

47. Averdunk, L. et al. Biallelic variants in CRIPT cause a Rothmund-Thomson-like syndrome with increased cellular senescence. Genet. Med. 25, 100836 (2023).

48. Oppermann, H. et al. CUX1-related neurodevelopmental disorder: deep insights into phenotype-genotype spectrum and underlying pathology. Eur. J. Hum. Genet. (2023) doi:10.1038/s41431-023-01445-2.

49. Schmetz, A. et al. Delineation of the adult phenotype of Coffin-Siris syndrome in 35 individuals. Hum. Genet. 143, 71–84 (2024).

50. Küry, S. et al. Unveiling the crucial neuronal role of the proteasomal ATPase subunit gene PSMC5 in neurodevelopmental proteasomopathies. medRxiv (2024) doi:10.1101/2024.01.13.24301174.

51. Li, D. et al. Spliceosome malfunction causes neurodevelopmental disorders with overlapping features. J. Clin. Invest. 134, (2024).

52. Rigter, P. M. F. et al. Role of CAMK2D in neurodevelopment and associated conditions. Am. J. Hum. Genet. 111, 364–382 (2024).

53. Laugwitz, L. et al. ZSCAN10 deficiency causes a neurodevelopmental disorder with characteristic oto-facial malformations. Brain (2024) doi:10.1093/brain/awae058.

54. Clark, T., Ciccarese, P. N. & Goble, C. A. Micropublications: a semantic model for claims, evidence, arguments and annotations in biomedical communications. J. Biomed. Semantics 5, 28 (2014).

55. Raciti, D., Yook, K., Harris, T. W., Schedl, T. & Sternberg, P. W. Micropublication: incentivizing community curation and placing unpublished data into the public domain. Database 2018, (2018).

56. Liu, J. et al. Natural History and Real-World Data in Rare Diseases: Applications, Limitations, and Future Perspectives. J. Clin. Pharmacol. 62 Suppl 2, S38–S55 (2022).

57. European Union. Charter of Fundamental Rights of the European Union, 2016. EUR-Lex. https://eur-lex.europa.eu/legal-content/EN/TXT/?uri=celex%3A12016P%2FTXT (2016).

58. Regulation (EU) 2017/745 of the European Parliament and of the Council of 5 April 2017 on medical devices, amending Directive 2001/83/EC, Regulation (EC) No 178/2002 and Regulation (EC) No 1223/2009 and repealing Council Directives 90/385/EEC and 93/42/EEC. https://eur-lex.europa.eu/legal-content/EN/TXT/?uri=CELEX%3A32017R0745.

59. Regulation (EU) 2016/679 of the European Parliament and of the Council of 27 April 2016 on the protection of natural persons with regard to the processing of personal data and on the free movement of such data, and repealing Directive 95/46/EC (General Data Protection Regulation). https://eur-lex.europa.eu/legal-content/EN/TXT/?uri=CELEX:32016R0679.

60. Deng, J. et al. ImageNet: A large-scale hierarchical image database. in 2009 IEEE Conference on Computer Vision and Pattern Recognition 248–255 (2009).

61. Huang, G. B., Ramesh, M., Berg, T. & Learned-Miller, E. Labeled Faces in the Wild: A Database for Studying Face Recognition in Unconstrained Environments. http://vis-www.cs.umass.edu/lfw/. (2007).

62. Boyadjiev, S. A. & Jabs, E. W. Online Mendelian Inheritance in Man (OMIM) as a knowledgebase for human developmental disorders. Clin. Genet. 57, 253–266 (2000).

63. den Dunnen, J. T. et al. HGVS Recommendations for the Description of Sequence Variants: 2016 Update. Hum. Mutat. 37, 564–569 (2016).

64. Stevens-Kroef, M., Simons, A., Rack, K. & Hastings, R. J. Cytogenetic Nomenclature and Reporting. in Cancer Cytogenetics: Methods and Protocols (ed. Wan, T. S. K.) 303–309 (Springer New York, New York, NY, 2017).

65. Kaye, J. et al. Dynamic consent: a patient interface for twenty-first century research networks. Eur. J. Hum. Genet. 23, 141–146 (2015).

66. Nellåker, C. et al. Enabling Global Clinical Collaborations on Identifiable Patient Data: The Minerva Initiative. Front. Genet. 10, 611 (2019).

67. Schoeman, L., Honey, E. M., Malherbe, H. & Coetzee, V. Parents’ perspectives on the use of children’s facial images for research and diagnosis: a survey. J. Community Genet. 13, 641–654 (2022).

68. Schmidt, A. et al. Next-generation phenotyping integrated in a national framework for patients with ultra-rare disorders improves genetic diagnostics and yields new molecular findings. medRxiv 2023.04.19.23288824 (2023) doi:10.1101/2023.04.19.23288824.

