## Supplementary Materials for "GestaltMatcher Database - A global reference for facial phenotypic variability in rare human diseases"

### Main components of the GestaltMatcher Database (GMDB) online platform

#### Gallery view

The Gallery view displays all GMDB-FAIR (Findable, Accessible, Interoperable, Reusable) case report images, which can be scrolled endlessly. It also enables clinicians to search for a gene or disorder of interest easily and allows for immediate visualization of all relevant portrait images (at-a-glance). Moreover, the user can search for Human Phenotype Ontology (HPO) terms or a specific publication's PubMed ID and Digital Object Identifier (DOI).

#### Research platform

The Research Platform is a section of the GMDB that allows clinician scientists to analyze the facial similarity of their cohorts using GestaltMatcher Artificial Intelligen (AI). To perform an analysis, a clinician scientist can select all FAIR cases of interest from the Gallery. It is also possible to add their own privately uploaded cases for analysis. The results of this analysis are displayed in a Pairwise Rank and Pairwise Distance Matrix, or t-SNE[1] projection, to explore the location of the analyzed patients in the Clinical Face Phenotype Space[2]. This information can then be used, for example, to increase knowledge of the facial phenotypic variability of a cohort, phenotype-genotype correlations, or lumping and splitting analysis.

### Special features of GMDB

##### Example for a micro-publication of a case report

As an incentive for clinicians to share patient data and help build a more diverse dataset, the GMDB allows collaborating clinicians to assign a DOI to their cases. The DOI transforms the case into a micro-publication. For this purpose, the case is also assigned a title and an Abstract. This information (title, abstract, DOI) is displayed on the landing page of a case (Supplementary Figure 2) and is available publicly without access to the GMDB. To protect the patient's data, phenotype and image data are only accessible after registration to the GMDB. For registered users, all details on the case can be seen, including all FAIR data and metadata (Supplementary Figure 1). Over the last two years from 2022 to 2024, this approach enabled the publication in GMDB of 2,312 images of cases that have not been published elsewhere.

One example of a published case with an assigned DOI concerns an individual of admixed ancestry (Gambian and European) who presented as an atypical case of Noonan syndrome. The patient had multiple lentigines and atypical facial dysmorphic features (hypotelorism and upslanting palpebral fissures instead of hypertelorism and downslanting palpebral fissures). These features had led the clinician to exclude a diagnosis of Noonan syndrome, only for it to be established later by exome sequencing. Interestingly, the analysis with GestaltMatcher identified a match with another Noonan syndrome patient as the 12$^{th}$ most similar case.

This case report was uploaded to the GMDB as a FAIR case and was assigned a DOI to enable it to be shared with the clinical and scientific community (Figure 3a). Metadata, multimodal data (profile images, skin, hand, and feet), and longitudinal data were subsequently added to this case (Figure 3a). As this example is a FAIR case, it can also be seen in the Gallery view (Figure 3b) and used for cohort analysis in the research platform to perform similarity comparisons (Figure 3c).

#### Digital Consent

##### Digital Consent for patient recruitment

To make the patient recruitment process as fast as possible and to optimally involve patients in this process, the option of using digital consent for informed consent for the project was integrated into the GMDB. This option can be an alternative to printed consent if the patients prefer it. However, digital consent can shorten the recruitment process, especially for retrospective recruitment, as a personal re-consultation with the patient is not necessarily required. It also enables patients to enter their own data at any time and upload their own images and files, which is particularly helpful if the attending physician does not have all the previous laboratory findings yet.

##### Usage of digital consent

For using digital consent, the clinician first needs to first create a case by clicking 'new patient' (Supplementary Figure 5a). In the next step, it must be indicated what kind of consent the patient should sign (at this time, we offer English and German) and a pseudonym (Supplementary Figure 5b). Afterwards, one must click "create patient" (Supplementary Figure 5c). Now, the case is created, and an invite can be sent to the patients. This can be done by sending out a prewritten email ('invite by mail') or by just copying a link ('invite by link') and sending this via mail or other medium preferred by the patient. During a patient consultation, it is also possible to sign the consent form directly e. g. on a tablet ('Sign consent') (Supplementary Figure 5d).

After the patient follows the link, he or she needs to create an account first, then the study information and consent form will be displayed (Supplementary Figure 6). After the patient signs the consent form, he or she can see his or her own data sheet and upload images and files. At the same time, the clinician will receive a notification to check the consent and uploaded files. After the clinician has checked what the patient has indicated in the consent, whether he or she has signed legally, and whether the privacy settings have been entered by the patient for each image or document as specified by him or her in the consent, the clinician can also make entries in the data sheet and upload files. After a final check of all details and privacy settings and the ranking of the distinctiveness for each uploaded image, the clinician can select the image to be displayed in the Gallery and then tick the 'consent obtained' box. Only after ticking this box will a FAIR case be visible for all GMDB users in the Gallery and usable for other scientists. A patient can access the data at any time via his or her account, upload new data or revoke consent. In the future, dynamic consent will also be implemented.

#### GMDB datasets

To enable transparency and reproducibility, we only used GMDB-FAIR for all the analyses in this study. However, in GMDB patient recruitment, we distinguished between the GMDB-FAIR dataset and the GMDB-private set (Supplementary Figure 7, Supplementary Figure 8). We introduced this distinction because it is known that patient consent to data sharing is higher when not shared with a broad mass, but only for a specific study[3]. However, many patients agree to controlled access for the general scientific community to advance research[3]. For this reason, patients can decide whether they agree to be part of the FAIR dataset or only want to be part of the GMDB-private set for GestaltMatcher AI training.

**GMDB-FAIR dataset**

This dataset is explained in detail in the main manuscript and the basis for all analyses in this paper (Supplementary Figure 7b).

**GMDB-Private dataset**

The private dataset (Supplementary Figure 7a) is only used for training and testing the GestaltMatcher AI. These cases involve patients who have only provided consent for the ‘private’ utilization of their data. Data is uploaded by the attending clinician or by the patient himself after he or she has received an invitation link to his or her case from the attending clinician. Consequently, while the case remains visible only to the uploading clinician, it is inaccessible to other users of the online platform and cannot be disseminated to external groups. It is also possible to upload only individual images or documents 'privately' to a FAIR case. This private dataset is used exclusively for the further training and improvement of the GestaltMatcher AI. However, the uploading clinician can also perform analyses in the research platform with this case and perform facial similarity comparisons.

### Supplementary Figures

**Supplementary Figure 1: Patient page for an individual with Leopard syndrome.** Each case report in the database has its own data sheet in which all image and metadata are stored in a clearly structured manner. It contains general demographic information such as gender and ethnicity, but also the uploading clinician, as well as a GestaltMatcher Database (GMDB) Case ID and a pseudonym that the uploading clinician can assign to the case. Either the informed consent has already been obtained in advance in paper form, which must be confirmed by the clinician at this point, or the patient has signed a digital consent, which will be indicated here. In addition, the information on the underlying disease with a link to the webservice Online Mendelian Inheritance in Man (OMIM) and the molecular information with disease gene, test method, and mutation in Human Genome Variation Society (HGVS) nomenclature format can be found. For each uploaded image, the image type and the person's age at the time the image was taken can be specified. In addition, for each image, there is information on whether the image can be shared with the GMDB community (Private - No (N)) or whether it should only be uploaded privately (Private - Yes (Y)). Further phenotypic information is documented using the Human Phenotype Ontology (HPO).

**Supplementary Figure 2: Landing page of the unpublished Noonan syndrome patient with an assigned DOI.** Each entry in the GestaltMatcher Database (GMDB) features a dedicated landing page tailored for non-registered visitors. This page presents information on the respective case, such as the Digital Object Identifier (DOI), the citation, the title, and an abstract. Additionally, it outlines the data that are accessible post-login in tabular form, including specifics on available image types related to the case, as is required by the metadata schema of DataCite. Details such as ancestry, gender, diagnosed disease, and the associated disease gene are visible on the landing page, while access to images and phenotypic information requires login. Upon login, users gain full access to the comprehensive data sheet of the case.

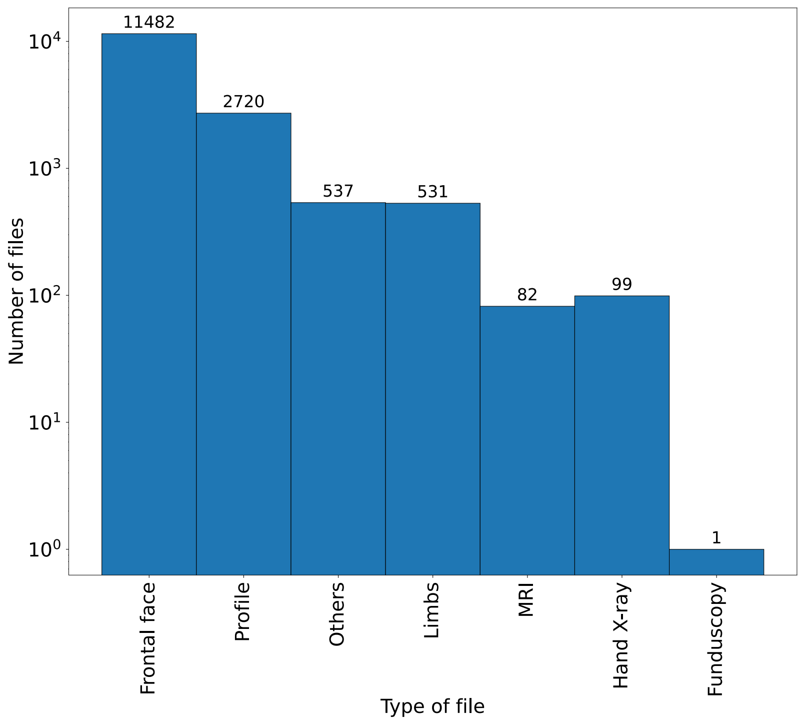


**Supplementary Figure 3: Overview of the image types in GestaltMatcher Database (GMDB). The x-axis is the file type, and the y-axis is the number of files for each category.** While the GMDB mainly consists of frontal facial images (11,482 in total), it is not limited to this type of image. It also includes a large number of profile images (2,720), as well as images of limbs, magnetic resonance imaging (MRI), hand x-rays, and funduscopy, among others.

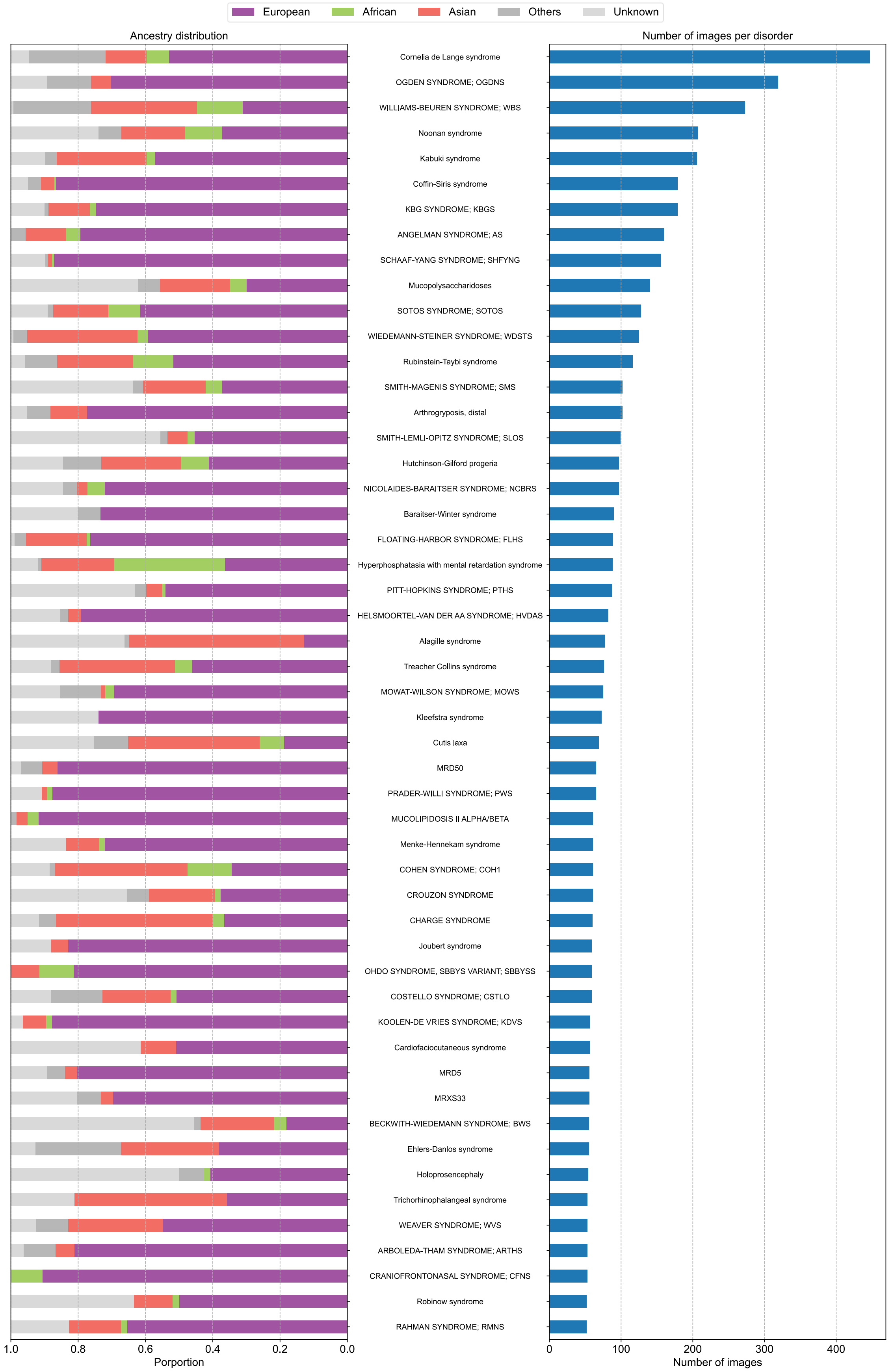


**Supplementary Figure 4: Each disorder's distribution of ancestries and image counts.** The left panel displays the distribution of the ancestries in the disorder, and the right panel shows the number of images in the disorder. The disorders with more than 50 images are shown in this figure.

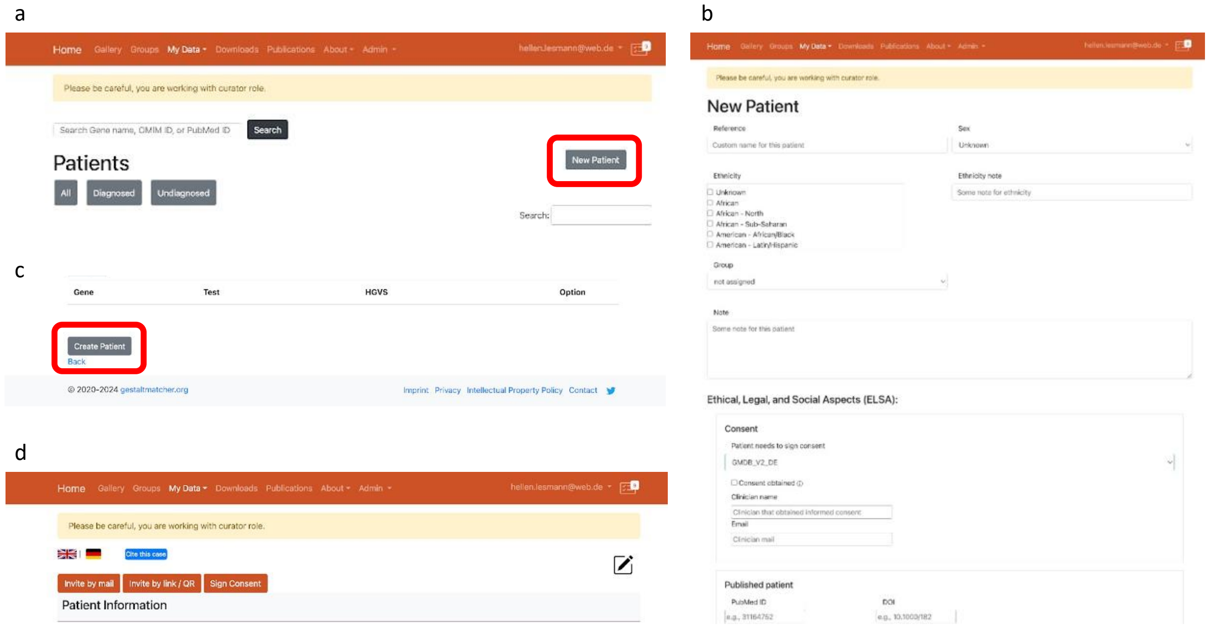


**Supplementary Figure 5: Creating a case and inviting a patient to GestaltMatcher Database (GMDB).** To invite a patient to the GMDB, a case must first be created. To do this, click on 'New Patient' **(a)**. You can then assign a pseudonym to this case and select which digital consent (English or German) should be displayed to the patient after registration **(b)**. Then click on 'Create Patient' to create the case sheet **(c)**. An invitation can then be sent to the patient **(d)**. There are 3 options for this: 1. 'invite by mail' - an e-mail text is automatically created for the patient, which contains a personal access link. 2. 'invite by link/QR' - the link is displayed, also in quick-response (QR) code form, and can be sent to the patient or can be scanned directly with a mobile device. 3. 'Sign consent' - the patient can sign the consent on a tablet directly during the conversation.

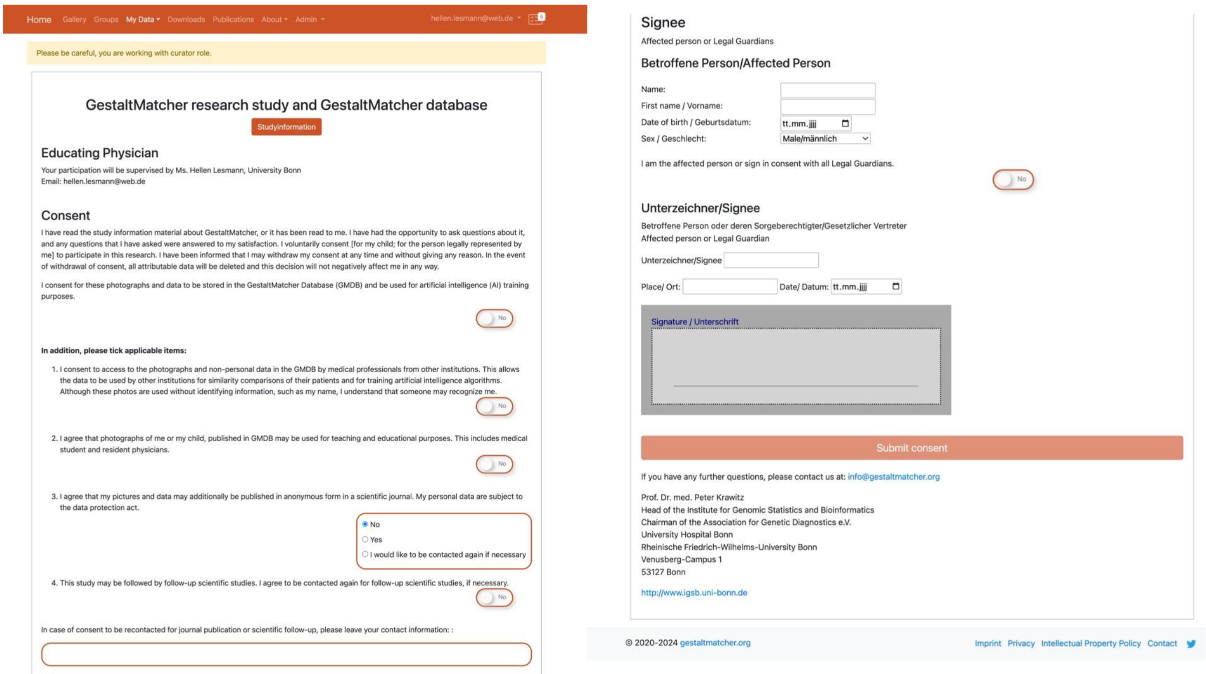


**Supplementary Figure 6: Digital consent.** The digital consent form is displayed to the patients after they register using their personal registration link. The entire GestaltMatcher study information can be viewed again under ‘study information’. The patients can then decide in detail which conditions of the study they agree to and which they do not. They can also enter their contact details if they agree to be re-contacted. At the end, they can sign digitally.

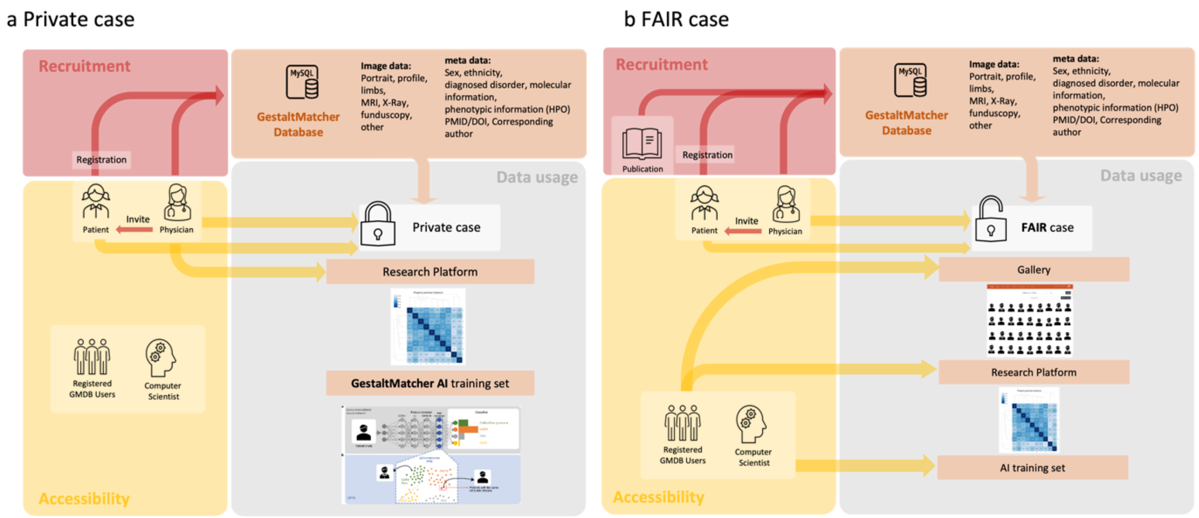


**Supplementary Figure 7: The different usages of Private case and FAIR case data. a)** A private case can be uploaded directly to the GestaltMatcher Database (GMDB) by a clinician or by a patient previously invited by the clinician. However, the data cannot be viewed by GMDB users. External computer scientists cannot use the data for their Artificial Intelligence (AI). Only the uploading clinician (and the patient) can view the case and use it to perform analyses in the research platform. In addition, the case can be used by the GestaltMatcher team to train and test the GestaltMatcher AI. **b)** FAIR (Findable, Accessible, Interoperable, Reusable) case report data comes from the literature, from clinicians or from the patient previously invited by the clinician. They can be viewed and analyzed by the uploading clinician and the patient but are also visible to all GMDB users in the Gallery and can be used by researchers in the GMDB for cohort analyses within the Research platform. After applying for an Institutional Review Board (IRB) approved study proposal, our board decides whether we should share the data and metadata with other researchers to train their AI.

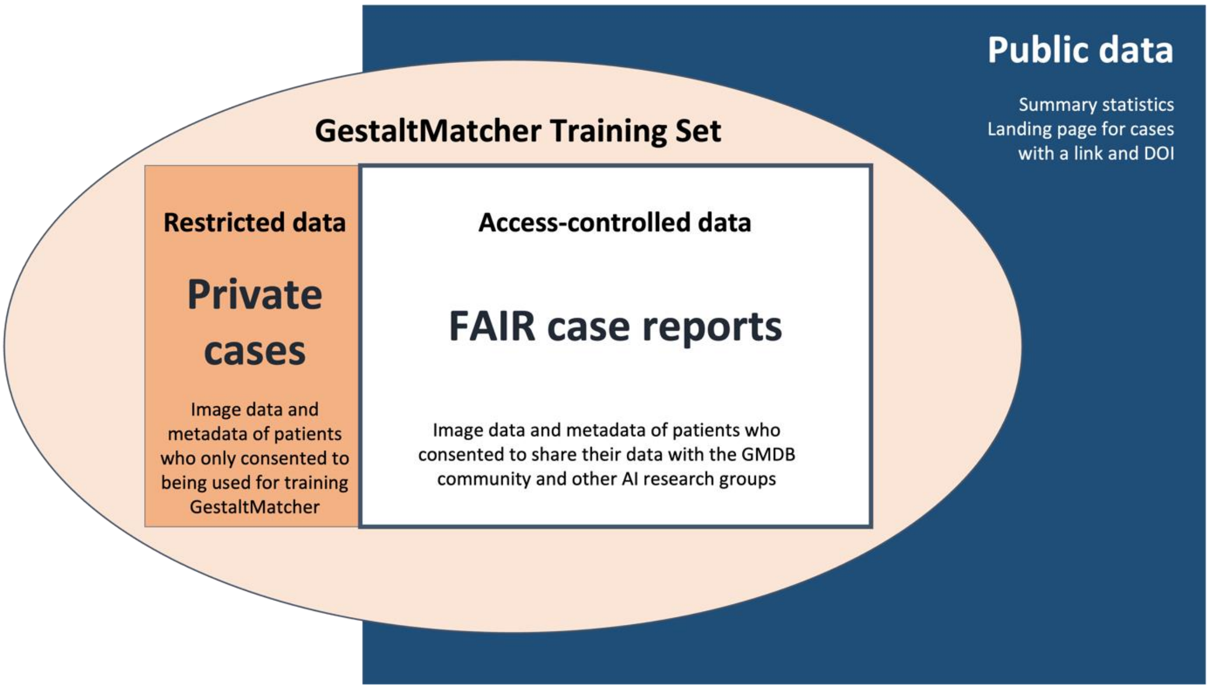


**Supplementary Figure 8: The GMDB training set consists of GMDB-private and GMDB-FAIR sets.** The GestaltMatcher Database (GMDB) contains two different datasets available on three different access levels. The GestaltMatcher training set consists of the private cases and the FAIR (Findable, Accessible, Interoperable, Reusable) case reports. The Private Cases cannot be viewed by GMDB users and are not shared with other researchers. They are only used to train the GestaltMatcher Artificial Intelligence (AI) (restricted data). The FAIR cases are subject to GMDB access control, only registered users can access them (access-controlled data). However, they can also be used by other external scientists to train their AI, as long as their application has been accepted by our Board and IRB approval has been obtained. On our website, we publish summarized statistics, and the landing page for cases with a Digital Object Identifier (DOI) can be viewed without login (public data).

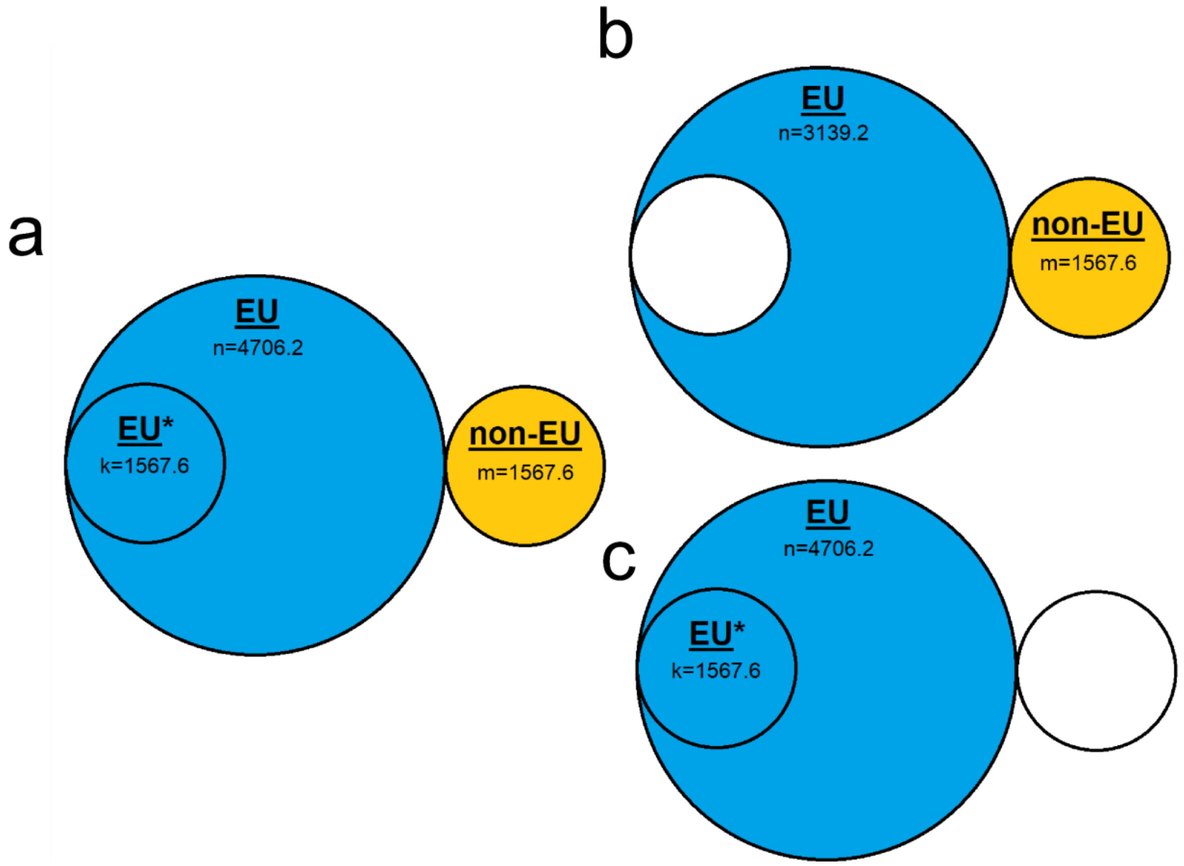


**Supplementary Figure 9: Illustration of training data used in ancestral analysis.** **a)** all suitable training data; **b)** EU + non-EU training data, selecting part of the EU set, which together with the non-EU set makes up the same size as EU + EU*; **c)** EU + EU* training data, selecting only EU training data, including the same base set as EU + non-EU.

#### References

1. van der Maaten, L. & Hinton, G. Visualizing Data using t-SNE. *J. Mach. Learn. Res.* **9**, 2579–2605 (2008).
2. Hsieh, T.-C. *et al.* GestaltMatcher facilitates rare disease matching using facial phenotype descriptors. *Nat. Genet.* **54**, 349–357 (2022).
3. Schoeman, L., Honey, E. M., Malherbe, H. & Coetzee, V. Parents' perspectives on the use of children's facial images for research and diagnosis: a survey. *J. Community Genet.* **13**, 641–654 (2022).
